# Springing Forward and Falling Back on Health: The Effect of Daylight Saving Time on Acute Myocardial Infarction

**DOI:** 10.1101/2022.07.06.22277274

**Authors:** Shinsuke Tanaka, Hideto Koizumi

**Author notes:** ^†^Corresponding author. Tufts University. We thank Hunt Allcott, Nathan Barker, Michael Boozer, Ellen Degnan, Laura Gee, Hugo Gerard, Meredith Houck, Michael Klein, Kazuki Motohashi, Muthoni Ngatia, Pace Phillips, Masayuki Sawada, Matthew D. Shapiro, Glynis Startz, Adam Storeygard, Tavneet Suri, Andreas Tiemann, Kosuke Uetake, Matthew White, Karen Zhang, Nicolas R. Ziebarth, and seminar participants at the 2015 ASSA-AEA, 6th Biennial Conference of the American Society of Health Economists, Osaka University, Tsukuba University, and University of Tokyo, and 2020 AERE-EEA for helpful comments. We also gratefully acknowledge the Indiana State Department of Health for generously providing data and Michele Starkey for her assistance with the data. Research results and conclusions expressed in this study are those of authors and do not necessarily reflect the views of the Indiana State Department of Health. All remaining errors are our own.

## Abstract

We examine the effects of Daylight Saving Time (DST) on the incidence of acute myocardial infarction (AMI) over three distinct time frames: short, medium, and long run. By exploiting the unique circumstances in Indiana, our findings highlight substantial short-term costs of increased AMI admissions *at* the spring transition by 27.2%, which last for approximately two weeks, are not displaced by counteractive reductions *during* the DST period, and are incurred at each transition *over* the years studied without adaptation. Together, in the context of current policy debates, these findings support terminating time adjustments yet provide little evidence to support permanent DST.

## I. Introduction

As of this writing, the U.S. Congress is currently considering legislation to make Daylight Saving Time (DST) permanent starting in 2023. DST is a common energy policy around the globe. Approximately one quarter of the world’s population is exposed to the semiannual ritual of adjusting the clock by an hour forward in spring (commonly known as “springing forward”) and by an hour backward in autumn (“falling back”). Its practice, however, has almost always been fraught with controversy and unpopularity.^1^ Recent compelling evidence has shown that the policy’s original intention to conserve energy by utilizing daylight in summer has backfired in that DST actually *increases* energy consumption.^2^ In addition, sizable evidence has substantiated adverse consequences on various economic activities as well as on the population where DST is practiced.^3^

But perhaps the most troubling concern over the practice of DST is that the seemingly-small yet abrupt one-hour change in how clocks are set can cause substantial anomalies to human health. When announcing the abolition of DST in Russia, former Russian President Medvedev succinctly summed up these anomalies by saying “Our biorhythms are damaged” (NEWSRT 2011). However, despite the widely accepted perceptions that DST leads to “fall back” in health outcomes, the direct linkage between the two has remained thin and inconclusive to date (Manfredini et al. 2019). Further, existing studies regarding the effects of DST on AMI exclusively focus on the transitional periods, whereas two important gaps remain in our understanding of i) the extent of the overall effects of DST during the DST period relative to the short-term effects of time adjustments and ii) whether people newly subject to DST adapt to the short-term effects of time adjustments at each DST transition over a period of years.

In this paper, we attempt to answer these long-standing questions by examining a relationship between DST and acute myocardial infarction (AMI), or heart attack, one of the primary health concerns associated with DST.^4^ Sleep deprivation (at the spring transition) and disrupted biorhythms (present at both transitions) are two physiological and pathological mechanisms through which DST can trigger AMI. We test whether DST increases the incidence of AMI over three distinct time frames: i) *at* the transitions in the short run; ii) *during* the overall DST period in the medium run; and iii) at each transition *over* a period of years in the long run.

To analyze the effects of DST in three temporal dimensions, we take advantage of the discrete nature of DST transitions and historical disputes over the adoption of DST in Indiana, which exempted a large majority of counties from adopting DST until 2006, resulting in county-date-year level variation in the practice of DST.^5^ This gives us three main advantages over the existing literature. First, to the best of our knowledge, this is the first paper to estimate the causal impact of DST on AMI by explicitly controlling for the seasonality effects at the transitional periods. Existing studies typically follow the time-series patterns of outcomes in days shifting to and from DST. These estimations are in principle similar to the regression discontinuity (RD) design, which compares mean outcomes across the discrete change in time on a certain day. An important concern with this and similar approaches that predominate in the relevant literature is that unobserved seasonality effects may bias the estimated treatment effects. Because daylight is known to serve as a zeitgeber (timekeeper) that can exactly synchronize or otherwise reset the biological clock within the human body, misalignment between the biological clock and the external environment at the seasonal transitions, even without DST transitions, is likely to cause disruption in biorhythms and increase the incidence of AMI.^6^ Our quasi-experimental research design offers a rare opportunity to address this concern by adopting the regression discontinuity difference-in-differences (RDDD) approach that differences out seasonality effects by using observations over the DST transitions when DST was not in effect.

Second, existing evidence regarding the relationship between DST and AMI exclusively focuses on the transitional periods. However, the short-term effects alone potentially distort the overall cost-benefit analysis of DST when the medium-term benefits are not accounted for. For example, the increased AMI cases at the transitions may reflect temporal displacement if the time change hastens heart attacks that would have occurred later even without the time change. Alternatively, the overall effects of DST on AMI may be beneficial if people utilize the longer daylight hours to engage in more physical activities. Our research design provides a rare context whereby we can address important gaps that remain in our understanding of i) the duration of the effects of the time adjustments, and ii) the extent of the overall effects of DST during the DST period.

Third, we present the first findings regarding whether those newly subject to DST continue to experience the short-term effects at each DST transition over a period of years, and thus whether there is adaptation to the effects of time change. For example, people newly subject to DST may begin to adjust their behaviors, e.g., by going to bed earlier on the night before the spring transition, thereby inflating the short-term effects in the first year of practicing DST while gradually or quickly lessening those effects over subsequent years.

Using administrative data on all AMI cases in Indiana in 2002–2012, our findings with respect to each of the three dimensions are as follows. First, the spring transition to DST increased AMI admissions by approximately 27.2%, whereas the smaller in magnitude, yet statistically significant, increase in AMI admissions after the autumn transition is likely to reflect the effects of seasonality, given the similar, often greater, increase in AMI admissions at the autumn transition, even when DST was not in effect. Second, while the effects of DST on AMI last for approximately two weeks after the spring transition, we find no evidence of temporal displacement by counteractive reductions in AMI admissions later, resulting in overall increased AMI admissions. Third, we find little evidence of adaptation over years; while the effect appears to be strongest in the first year in which populations are subject to the spring transition, we find no qualitatively or statistically significant decline in the effects of the spring transition over our study period. Together, these findings hold important policy implications in the context of current policy debates, providing strong support for terminating time adjustments yet no evidence to support permanent DST in terms of health benefits.

## II. Background

### A. Medical Linkage between DST and AMI**^7^**

Two channels explain the physiological and pathological mechanisms through which DST triggers AMI. First, DST directly disrupts circadian rhythms, an intrinsic 24-hour clock that synchronizes with external environmental factors such as daylight duration. An increasing number of studies in the medical literature document the close relationship between circadian rhythmicity and the cardiovascular system. Second, DST, directly and indirectly through disrupted circadian rhythm, can harm the quality and duration of sleep, which triggers cardiovascular disease. Using data from the American Time Use Survey, Barnes and Wagner (2009) show that the average sleep duration is 40 minutes shorter on the night of the spring transition, whereas no more sleep is gained at the autumn transition despite an extra hour on the clock.^8^

Although these studies illustrate plausible channels through which DST may cause AMI, the medical literature documenting the direct association between the two is relatively scarce and, when it does exist, has generated little consensus, as summarized in the recent meta-analysis study by Manfredini et al. (2019).^9^ Importantly, existing studies regarding the effects of DST on AMI exclusively focus on the transitional periods, e.g., only the first week after the time adjustment, whereas our study addresses important gaps in our understanding of: i) the duration of the effects of the time adjustments, ii) the extent of the overall effects of DST during the DST period relative to the short-term effects, and iii) whether people newly subject to DST adapt to the short-term effects of time adjustments at each DST transition over a period of years.

### B. Historical Debates over DST in Indiana**^10^**

Internal disputes over the practice of DST rendered Indiana a special case with multiple time zones and exemption from DST. As a result, during the period between 1991 and 2005, 15 out of 92 counties observed DST, while another 77 counties did not (Online Appendix Figure B1). In 2005, the Indiana state legislature passed a law that required all counties to observe DST starting in 2006.^11^ For the purpose of identifying the effect of DST on health, it is worth noting that the variation in time zone and related DST observance status in Indiana is based on the “convenience of commerce,” as evaluated by the Department of Transportation, and thus unrelated to other factors such as health concerns.

## III. Data

The primary data for our analysis comes from nonpublic administrative records from Indiana Hospital Discharge Data (Indiana Department of Health 2012) that cover all AMI cases in Indiana from 2002 through 2012.^12^ The data are collected and managed by the Indiana State Department of Health^13^, and by special arrangement we obtained access to patient-level observations. The data contains detailed information on patients, such as age, gender, county of hospital, admitted date, discharge date, type of insurance a patient possesses, and whether s/he is an inpatient or outpatient, total medical charges before insurance is applied, and length of hospital stay for inpatients. In total, we observe close to 185,000 hospital admissions which constitute one of the most comprehensive datasets ever applied to this topic. For the main analysis, we focus on hospital admissions on weekdays because AMI admissions are known to be distinctively lower on weekends (Witte et al. 2005); we then attest the robustness of the findings by including weekends. Online Appendix Table C1 presents the descriptive statistics for the sample used in the analysis: the working-age population aged between 20 and 64.

## IV. Econometric Framework

### A. Regression Discontinuity (RD) Methods

To estimate the effect of DST on AMI at the transitional period, we employ the regression discontinuity (RD) design by including a flexible function of the running variable, f(*·*);

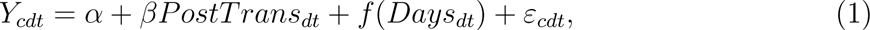

where *Y_cdt_* is the outcome of interest in county *c* on day *d* of the year *t*, *PostTrans* is a dummy variable for observations after the transition to or from DST, and *Days* denotes the number of days from the DST transition, normalized to zero on the first day of the transition, and thus *PostTrans* = 1(*Days ≥* 0). Our main outcome is AMI admission rates per one million working-age population.^14^ The parameter of interest, *β*, measures a discontinuous change in the outcome at the DST transition date, after flexibly controlling for the trends leading to and following to the transition.

We follow Imbens and Lemieux (2008) to use the uniform weight and specify the function to be a linear model whose slope is allowed to vary before and after the threshold:

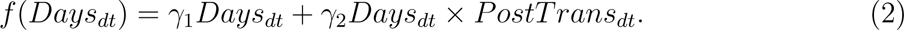

We further extend the model by including a full set of control variables

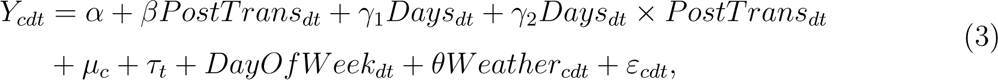

where county fixed effects *µ_c_* control for permanent county-level characteristics associated with variation in AMI incidence, year fixed effects *τ_t_* absorb overall time trends in AMI incidence common across counties within a year, *DayOf Week* is a set of day-of-week fixed effects, and *Weather_cdt_* controls for weather variability by including bins of every 3-degree Celsius interval and total precipitation at the county-daily level.^15^ The regression is weighted by the denominator of the outcome, e.g., the working-age population, as counties with larger populations provide more precise estimates in computing averages.^16^ The standard errors are clustered at the county level to correct heteroskedasticity in the error term and allow for correlation between observations within cluster. The selection of bandwidth provokes the known tradeoff between efficiency and bias. We begin with 4 weeks before and after the DST transition and test the robustness to various other bandwidths.^17^

The causal inference of Equation (3) is warranted if the conditional expectation function, *E*[*ε|Days*], is locally smooth over the DST transitions. That is, no factor affecting AMI other than DST exhibits a discontinuity at the DST transitions. However, such an assumption should not be taken for granted because the well-known circadian effect during the seasonal transitions is likely to confound the estimates. For example, changes in zeitgeber, such as the length of daytime and exposure to greater evening ambient light, disturbs an organism’s biological rhythm (Durgan and Young 2012; Burgess 2013) even in the absence of DST. Sinc the timing of DST is by design commensurate with seasonal transitions, due to its original intention to utilize longer daylight, the estimated DST effect is likely to spuriously pick up such seasonal effect.

### B. Regression Discontinuity Difference-in-Differences (RDDD) Methods

One of our main innovations over previous studies is that we can directly observe and control for seasonality effects, if any, based on the unique experiences in Indiana. Unlike most countries and those U.S. states that either have or have not observed DST over a long period of time, our research context offers a rare opportunity to directly test the identification assumption by applying the same model to observations when DST was not practiced. Because there was indeed no time change in those counties, the effect of DST must be zero. Any significant estimates would invalidate the RD assumption by indicating that there is a discontinuous trend in the outcome variable around the threshold for reasons other than DST. Further, we can estimate the true DST effect by differencing out the seasonality effect, if any, by using all observations regardless of the practice of DST and estimating the regression discontinuity difference-in-differences (RDDD) model:

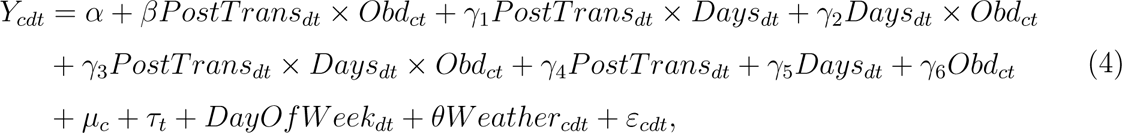

where the additionally introduced variable, *Obd*, is a dummy variable taking the value of one if the county observed DST in that year for that season.^18^ The basic intuition behind this model is that, to the extent there is an underlying discontinuity in AMI trend at transition dates even in the absence of transitions, we difference it out from the estimated treatment effect by using observations when DST was not practiced. The true effect of time change at the transition date, net of the seasonality effect, is captured by the coefficient *β*. The standard errors are clustered at the county level.

Because we directly control for discontinuities in the underlying conditional function at the threshold, we no longer require the usual continuity assumption of the conditional expectation function. Rather, the model assumption is that any changes in the outcome at the DST transition when DST is not practiced provide a counterfactual for what would have happened in the absence of DST for observations when DST was practiced. Thus, the identification assumption is that *E*[*ε|Days, Obd* = 1] = *E*[*ε|Days, Obd* = 0] holds at least locally across the DST transitions. With the model that includes the county-fixed effects, the estimated effect utilizes only the variation in DST practice by treatment counties where DST became in effect from 2006, and the counterfactual is provided by the same set of counties when DST was not in effect before 2006. We attest the validity of such identification assumption by applying the same RDDD model to the pre-reform period; if the assumption is valid, there should not be any effect on outcomes at the DST transitions in years before DST was adopted. The potential confounding variables in RDDD are those that discontinuously change at the DST transition only among the treatment counties and in the post-reform period. We are not aware of any variables that meet these conditions.

Further, we extend the model to explore the effects at every DST transition over the years studied to yield an inference regarding adaptation to the time adjustments among people who are newly subject to DST. In particular, we run the event-study RDDD model adapted to estimate the effects of the time adjustments in each year:

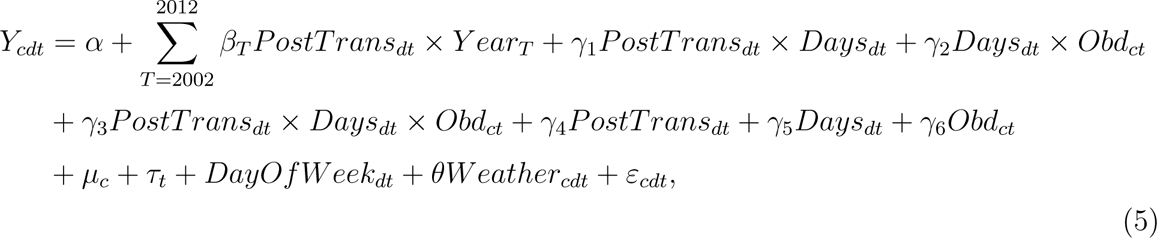

where *Y ear_T_* is an indicator variable for each year between 2002 and 2012. The omitted group is 2005, the year before the policy reform.^19^ Each *β_T_* indicates the effects of time adjustment in year *T* relative to that in 2005, and we expect the coefficients to be zero during the pre-reform period as a test of our identification assumption that within-county variation in AMI admissions at DST transitions over years is similar during the pre-reform period, whereas the coefficients during the post-reform period infer the rates of adaptation to the effects of time adjustments at every transition over a period of years.

### C. Difference-in-differences (DD) Methods

While the RD and RDDD analyses provided above are useful in estimating the causal effect of DST on AMI at the transitions, it is of limited use in estimating the medium-term effects of the time adjustments. In particular, we explore i) the duration of the effects of the time adjustments, and ii) the extent of the overall effects of DST during the DST period relative to the short-term adverse effects. An empirical investigation of these questions has been impeded by an empirical challenge to construct valid control observations during the DST period for what would happen in the treatment observations without a treatment. Our research design provides a rare context whereby we can estimate such medium-term effects by utilizing the unique policy change in Indiana.

To investigate the first question, we employ the DD model that explores the lasting effects of DST on AMI over several weeks after the transitions. In particular, using days surrounding the time adjustments, we estimate:

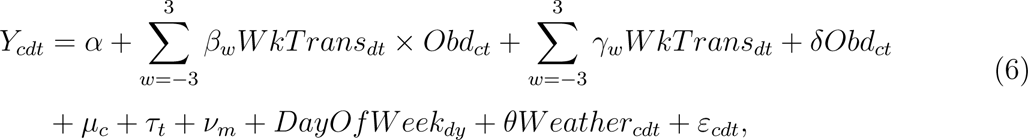

where *WkTrans* is a dummy variable for each week after the time transition, normalized to be zero in the week of the transition, and *ν_m_* is the month fixed effects. The weeks after the transitions are considered as the treatment group, whereas those before are the control group, and *Obd* is regarded as an indicator for the post-reform period. The parameters of interests are *β_w_*’s, which estimate the effects on AMI admissions in each week over the transitional period in years when DST was in effect relative to years when DST was not in effect. Thus, we expect the coefficients to be zero for weeks before the time transition, whereas the coefficients after the time transition indicate the lasting effects of the time adjustments. The regressions are weighted by the working-age population at the county level. The omitted group is the week before the time transition, e.g., *w* = *−*1.

Further, to investigate the second question, we compare the changes in AMI admission rates from pre- to post-reform periods among treatment counties where DST was not in effect before 2006 with those changes in control counties where DST was in effect throughout the study period using observations during the DST period:

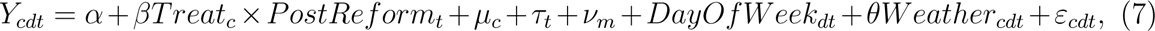

where *Treat* is an indicator variable taking the value of one for treatment counties where DST was not in effect before 2006, *PostReform* is an indicator variable taking the value of one for the post-reform period, e.g., post-2006 for most counties,^20^ while other variables are similarly defined as above. The coefficient of interest, *β*, measures the average effects of switching from Standard Time to DST over all days during the DST period, which captures the net effects of the spring transition and potential medium-term benefits of longer daylight. The regressions are weighted by the working-age population at the county level. All standard errors are clustered at the county level.

The model assumption of Equation 7 is that the treatment counties have similar trends to the control counties in the absence of treatment, e.g., the parallel trend assumption. We will present evidence to support such assumption by showing that the outcomes evolved similarly between the treatment and control observations during the pre-reform period. If any systematic differences in characteristics confounded our estimates, e.g., an age composition or underlying health behaviors, we would observe differential patterns in the pre-reform period as well. The potential confounding variables in DD are those that changed systematically only in the treatment counties in the same year as the policy changed. We are not aware of any variables that meet such condition.

## V. Results

### A. Short-term Effects at the Cutoffs

We begin with the graphical illustration of how DST transitions affected AMI admission rates.^21^ Figure 1 presents the trends of AMI admission rates for eight weeks before and after the DST transition. The *y*-axis measures the admission rates, residualized by the day-of-week effects, county fixed effects, year fixed effects, and temperature and precipitation effects, and *x*-axis measures the number of days after the transition, normalized to zero on the date of the DST transition. Panel A uses observations surrounding the spring transition when DST was in effect. We observe that while AMI admissions fall in the month before the spring transition, a sharp and discontinuous increase in AMI admissions is observed exactly at the beginning of DST. To test if seasonality effects can explain the increased AMI around this timing, Panel B plots an analogous figure using only the observations when DST was not in effect before 2006. It shows that there is no such discontinuity around the threshold for these observations; otherwise the trend appears to be similar to counties that observed DST.

**Figure 1:**
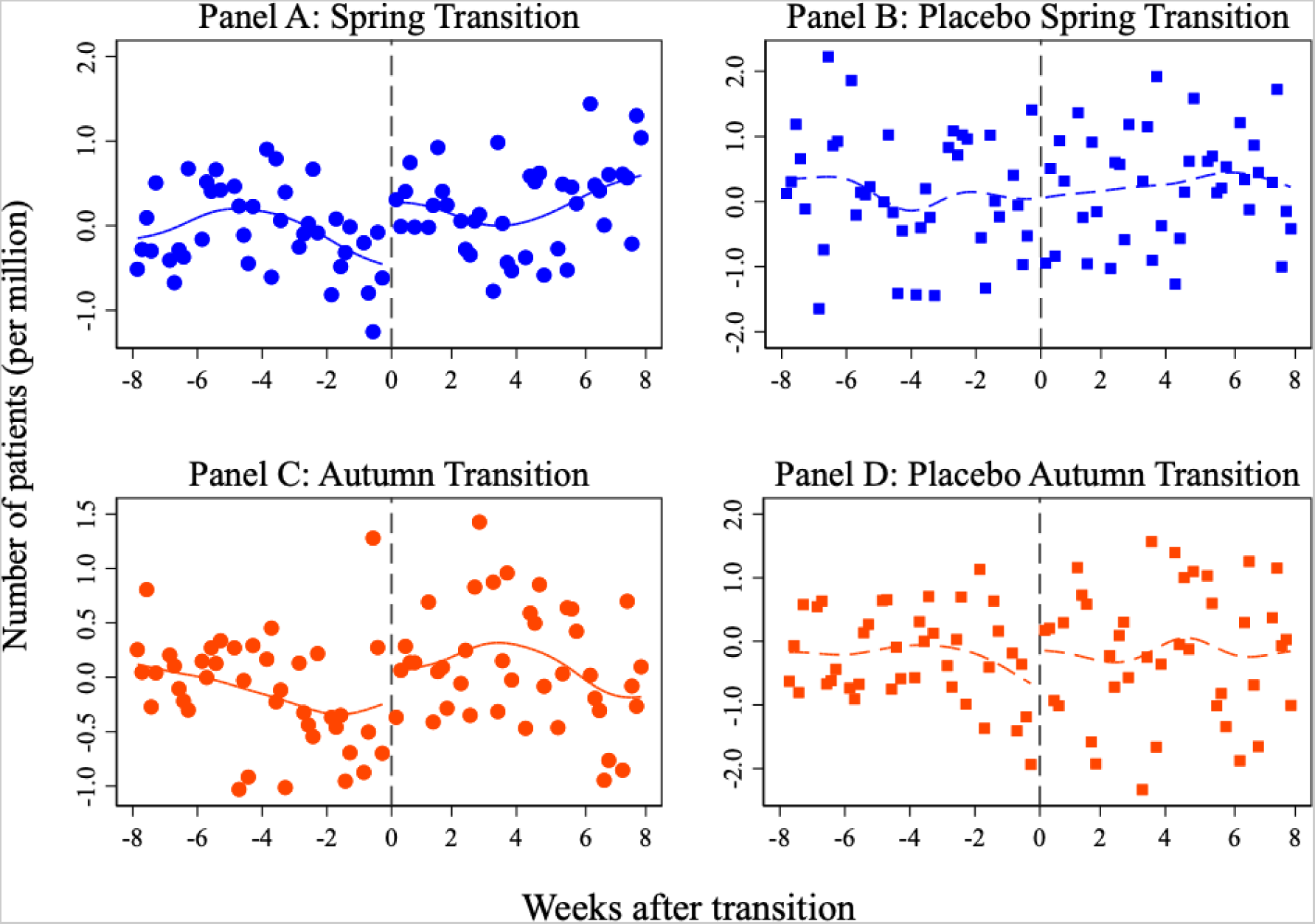
AMI Admissions around DST *Notes*: These figures plot the daily average AMI admission rates per one million working-age population among the working population aged between 20 and 64, weighted by the working population at the county level, residualized by the day-of-week effects, county fixed effects, year fixed effects, and temperature and precipitation effects. The lines indicate smoothed local polynomial trends with the bandwidth of seven days. Panels A and C include observations when the respective DST was practiced, whereas Panels B and D include observations when the respective DST was not practiced.

We now turn to the distinct observations for the autumn transition. On the one hand, Panel C illustrates a similar increase in AMI admissions at the DST transition for counties that practiced DST. On the other hand, Panel D highlights that AMI admissions also increased in counties that did not practice DST. Because there was no time shift for these observations, such an increase in AMI cannot be explained by DST and highlights the seasonality effect at the timing of the DST transition that triggers AMI even without DST.

The evidence above that placebo effects are present suggests that it is important to adequately control for the seasonality effect by constructing the appropriate counterfactual in order to estimate the true effect of DST, which is achieved by RDDD. Table 1 presents evidence based on the RDDD model as specified by Equation (4) using all samples, including both counties that did and did not practice DST, in which observations when DST was not practiced allow us to adequately control for the seasonality effects.^22^

**Table 1:** The RDDD Estimates of the Effects of DST Transition on AMI Admissions

|  | (1) | (2) | (3) | (4) |
| --- | --- | --- | --- | --- |
| *Panel A: Spring* |  |  |  |  |
| PostTrans×Obd. | 1.487***<br>(0.546) | 1.494***<br>(0.545) | 1.507***<br>(0.545) | 7.076**<br>(3.453) |
| Mean | 5.548 | 5.548 | 5.548 | 38.467 |
| N | 40,480 | 40,480 | 40,480 | 8,096 |
| *Panel B: Autumn* |  |  |  |  |
| PostTrans×Obd. | -0.468<br>(0.599) | -0.512<br>(0.599) | -0.533<br>(0.600) | -1.896<br>(3.219) |
| Mean | 5.393 | 5.393 | 5.393 | 36.811 |
| N | 38,824 | 38,824 | 38,824 | 8,096 |
| Fixed effects | N | N | Y | Y |
| Controls | N | Y | Y | Y |
| Level of obs. | Daily | Daily | Daily | Weekly |

*Notes:* This table presents the effects of DST on the daily AMI admission rates per one million working-age population at the county level in Columns (1)–(3) and the weekly AMI admission rates per one million working-age population at the county level in Column (4) among the working population aged between 20 and 64 based on the RDDD model of Equation (4). The treatment observations are the observations around the DST transition when DST was observed, whereas the control observations are analogous observations when DST was not observed. The bandwidth is 4 weeks before and after the transition. The regressions are weighted by the working population at the county level. Fixed effects include year- and county-fixed effects, and controls include the day-of-week effects, daily mean temperature with bins of every 3-degree Celsius, and daily total amount of precipitation. The mean value of the dependent variable is computed for the pre-transitional period when DST was practiced, weighted by the working population at the county level. The standard errors clustered at the county level are reported in the parentheses.
\*\*\* $p < 0.01$ \*\* $p < 0.05$

The findings are consistent with what is expected from the graphical results, suggesting that AMI cases increased after the spring transition but not after the autumn transition. Importantly, Column (3) controls for the county-fixed effects, and therefore counterfactual is constructed from their own counties when DST was not practiced before 2006. Overall, the preferred model in Column (3) suggests that AMI admission rates increased by 1.507 per one million working-age population (incidence ratio (IR) = 1.272, 95%CI [1.076 1.467], *p* = 0.007, *n* = 40,480) by moving into DST. In contrast, AMI admission rates decreased by 0.533 per one million working-age population (IR = 0.901 95%CI [0.68, 1.122], *p*=0.376, *n*=38,824) by moving out of DST, although the point estimate is not statistically significantly different from zero. Column (4) presents the consistent results by the week-level observations.^23^

Online Appendix D.3 attests the robustness of the RDDD results to various other specifications. Briefly summarizing the results, we find that the findings above are robust to: i) using the count model, such as poisson and negative binomial models, ii) using various optimum bandwidth selections, iii) including weekends, iv) using polynomial running variables, v) using alternative control groups, and vi) accounting for commuting effects. Further, the permutation test that assigns the placebo DST transition on each date outside the DST transitional period confirms that our estimated effect is clearly an outlier and has an implied *p*-value of 0.0068 (Online Appendix D.4). In addition, Online Appendix D.5 explores potential heterogeneities in the effects of DST on AMI across various dimensions by applying the main RDDD analysis to a set of subsamples. Briefly summarizing the results, while we find that the effects are relatively similar across various subgroups, if anything, the effects are stronger for women relative to men, people aged above 50 relative to below 50, and those with private insurance relative to those with Medicaid. Last, Online Appendix D.6 shows that including fatal incidences does not alter our conclusions.

### B. Medium-term Effects after Time Adjustments

This section explores the medium-run effects of time adjustments.^24^ Figure 2 presents the estimated coefficients based on Equation (6). We find that the coefficients are zeros in weeks leading to the time adjustments, confirming the identification assumption that the AMI admissions trended similarly before the spring transition between the pre- and post-reform periods. In contrast, consistent with the RDDD results, we find that the week after the spring transition experiences a substantial increase in AMI admissions. The size of the effect in the first week is similar to that predicted by RDDD analysis, suggesting that the impacts at the spring transition remain of similar magnitude throughout the first week (Online Appendix Table E1). More importantly, the results highlight slightly lower, yet similar, magnitudes of the effects in the second week after the spring transition. The point estimate halves in the third week and diminishes further to virtually zero in the subsequent week. These results suggest strongly that the effects of the spring transition last for two weeks after the time adjustment. Figure 1 also illustrates a consistent pattern of a discontinuous jump in AMI admissions at the spring transition that appears to persist for two weeks before reversing to essentially the same seasonal path observed before the transition.

Figure 3 presents the results from the event-study analysis using Equation (A.2) in Online Appendix E based on the DD model of Equation (7) that explores the overall effects of time adjustments during the DST or Standard Time period. Panel A presents the results during the DST period. We observe that AMI admission rates are virtually zero up to the year of treatment, ensuring that that there was no obvious pre-existing differential trend between the treatment and control counties; if any, AMI admissions were declining in the treatment counties. Yet, there is an evident increase in the first year subject to the spring transition, and we cannot reject that the effects are similar thereafter. Overall, the introduction of DST resulted in an increased daily AMI admission rates by 0.479 per one million working-age population (IR = 1.076, 95%CI [1.064 1.240], *p* = 0.090, *n* = 159,160) (Online Appendix Table E2). In contrast, consistent with the RDDD results, Panel B shows that AMI admission rates do not change after the autumn transition during the Standard Time (Online Appendix Table E2). These results suggest that the overall AMI admission rates increased during the DST period after the policy reform, and there is no evidence of any potential benefits of the longer daylight hours during the DST period that offset the short-term costs.^25^

**Figure 2:**
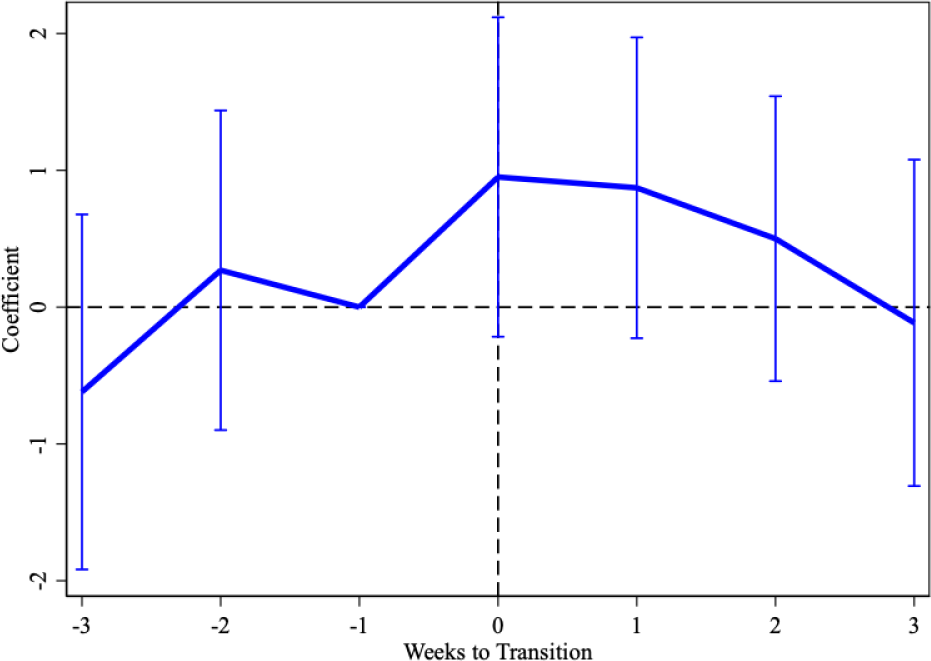
Lasting Effects of the Spring Transition *Notes*: This figure presents the effects of the spring transition on the AMI admission rates per one million working-age population over weeks before and after the time adjustment, normalized to zero in the week of the transition, based on Equation (6). The regressions are weighted by the working-age population at the county level. The vertical lines indicate the 95% confidence interval. The underlying estimates are presented in Online Appendix E.

**Figure 3:**
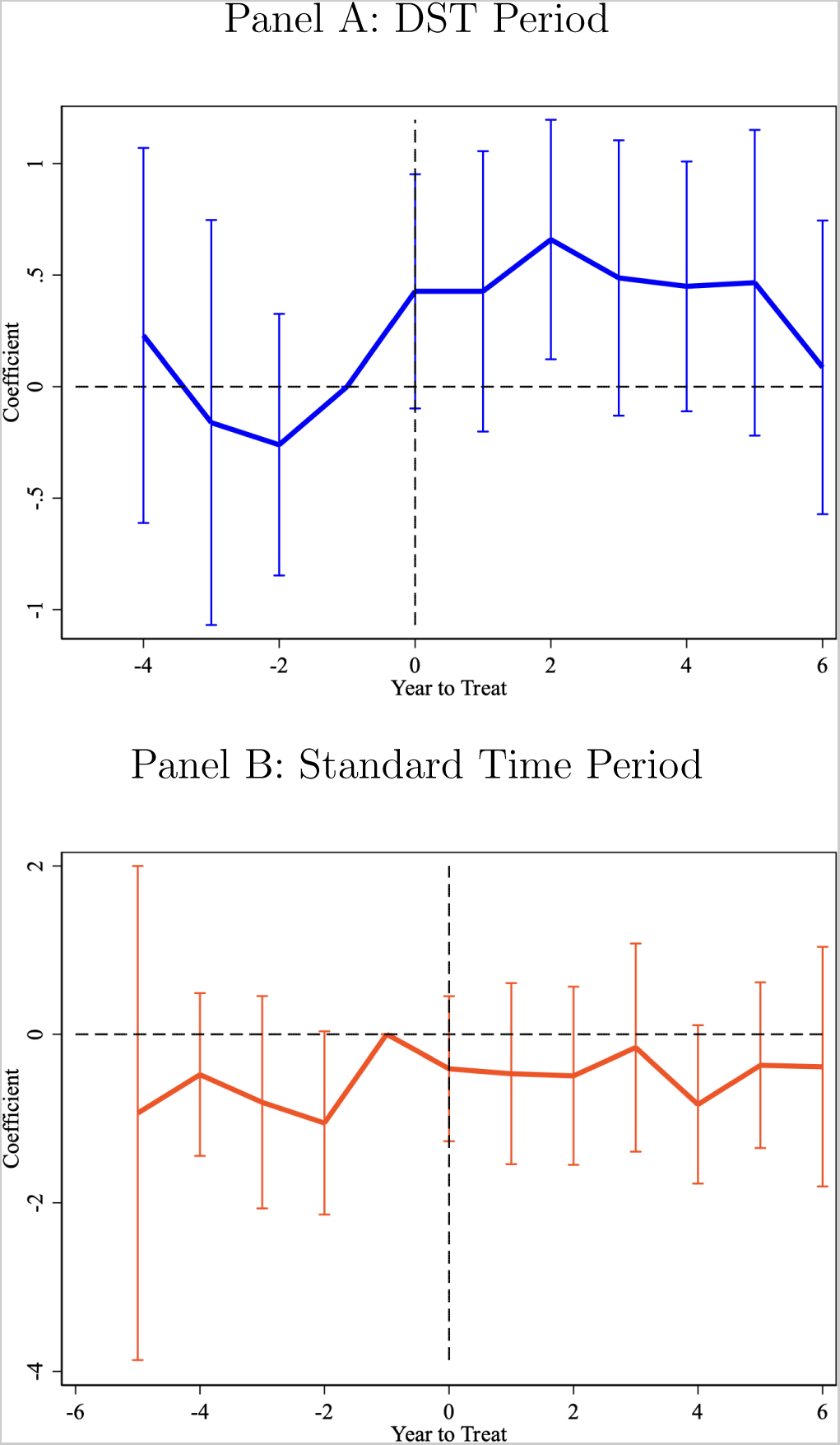
Effects of Time Adjustments during DST and Standard Time Periods *Notes*: This figure presents the effects of the time adjustments on AMI admission rates per one million working-age population during the DST period in Panel A and during the Standard Time period in Panel B. Each coefficient is estimated using the event-study model based on the DD model of Equation (7):

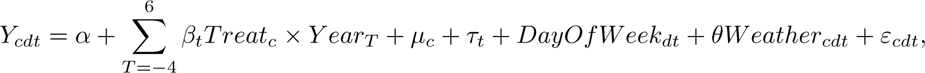

where the additional variable of *Y eart_T_* is an indicator variable for each number of years from the first year of practicing time adjustment, e.g., *T* = 0 in 2006 for most treatment counties. The omitted group is the year before the treatment starts, e.g., *T* = *−*1. The outcomes are the daily AMI admission rates per one million working-age population. The regressions are weighted by the working-age population at the county level. The vertical lines indicate the 95% confidence interval.

### C. Long-term Effects via Adaptation

Last, we are interested in exploring the effects of DST transitions at each transition over a period of years. In particular, we investigate whether people newly subject to DST adapt to the effects of time adjustments over time.

Figure 4 plots the estimated effects of the spring transition in each year based on Equation (5). The estimated coefficients are small and not statistically significantly different from zero over years before the policy reform, ensuring that there is no discontinuous change in AMI admissions at the spring transition when DST was not in effect. As aforementioned, this evidence supports the identification assumption of RDDD in that within-county variation in AMI admissions at the DST transitions across years is similar in the pre-reform period. In contrast, large and significant effects are observed starting in 2006. Notably, the estimated coefficient is the largest in 2006, and the estimated effects are high and consistent over the subsequent years studied.

**Figure 4:**
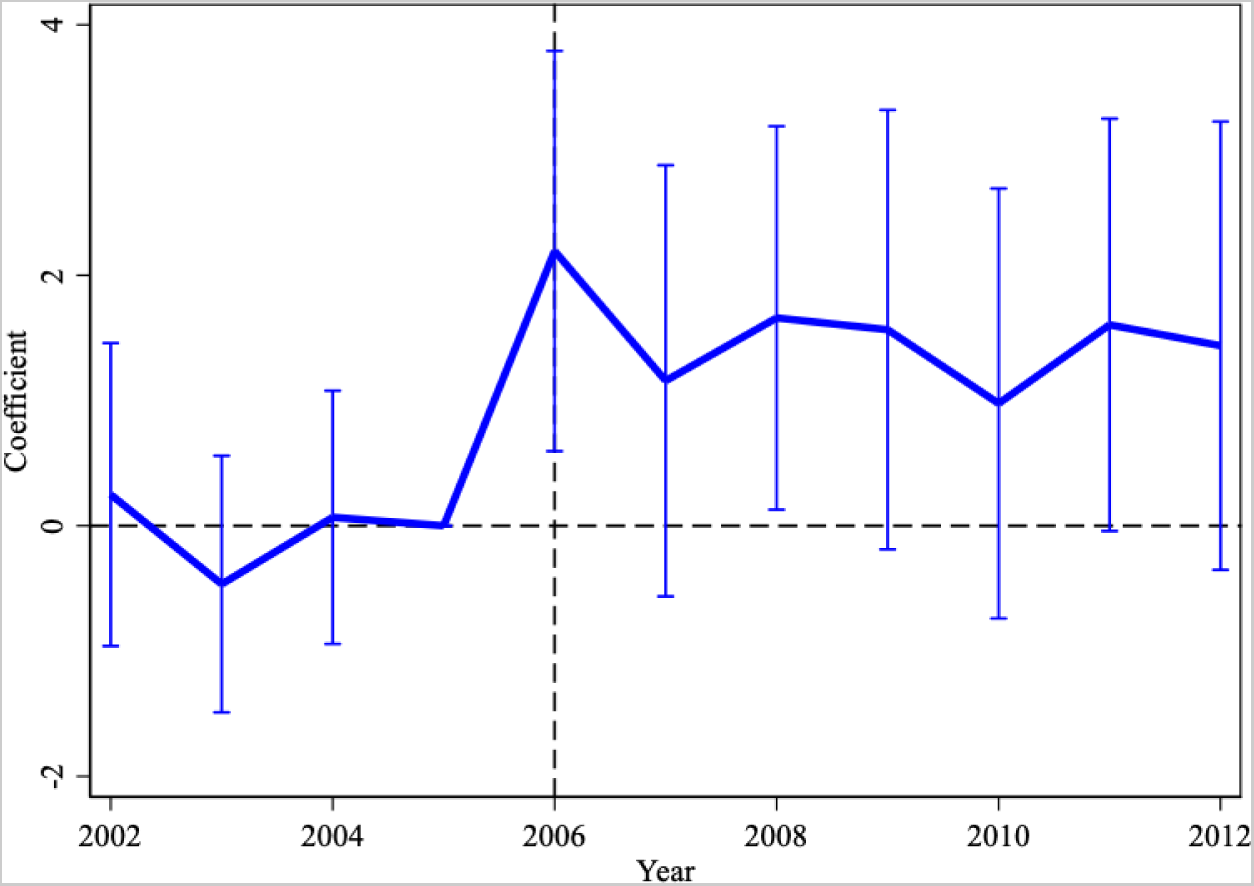
Effects of Spring Transition over Years *Notes*: This figure presents the effects of the spring transition on the AMI admission rates per one million working-age population in each year based on the RDDD model specified by Equation (5). The regressions are weighted by the working-age population at the county level. The vertical lines indicate the 95% confidence interval.

Overall, we find little evidence of adaptation; while the effect appears to be strongest in the first year subject to the spring transition, these results indicate no qualitatively or statistically significant decline in the effects of the spring transition at each transition over our study period.^26^

## VI. Conclusions

This paper presents empirical evidence regarding the effects of DST transitions on AMI admissions. We offer three main findings. First, DST increases AMI admissions only at the spring transition. While there appears to be a small increase in AMI occurring at the autumn transition, this increase becomes negligible after differencing out the seasonality effect. Second, while we find that the effects of DST last for approximately two weeks after the spring transition, we find no counteractive reduction in AMI admissions following the spring transition during the DST period, documenting no evidence of overall net benefits of longer daylight during the DST period. Last, such adverse impacts on health are high and persistent at each spring transition over a period of years without evidence of adaptation.

These findings hold important policy implications in the context of current policy debates by providing strong support for terminating the time change yet providing no evidence of net health benefits by permanent DST. The overall cost-benefit analysis of permanent DST needs to be weighted against benefits of longer daylight in other dimensions.

## Data Availability

The data used in the present study are proprietary.

## Online Appendix

### Overview

This Online Appendix provides additional information to what is already presented in the main text as well as additional evidence to attest the robustness of the findings in the main text to various other specifications and contexts. What follows is a brief summary of what each appendix presents.

Appendix A presents the detailed literature review regarding the medical linkage between DST and AMI.

Appendix B presents the detailed historical background over the practice of DST in Indiana.

Appendix C presents additional information on the data and variable constructions used for the main analysis. Further, it provides the summary statistics for the sample used in the main analysis.

Appendix D presents additional results on the short-term effects of DST at the transitional periods. Specifically, we present the results based on the RD model, the results using the weekly observations, the results from the robustness checks, the results from the permutation test, and the heterogenous treatment effects of DST across various subsamples.

Appendix E presents additional results for the medium-term effects of time adjustments during the DST and Standard Time periods.

Appendix F presents additional results regarding the long-term effects of time adjustments at each transition over a period of years.

## A. Medical Linkage between DST and AMI

Figure A1 illustrates several pathological and physiological mechanisms through which DST triggers AMI. Two channels explain the physiological and pathological mechanisms through which DST triggers AMI.

First, DST directly disrupts circadian rhythms, an intrinsic 24-hour clock that synchronizes with external environmental factors such as daylight duration.^1^ The 24-hour biological rhythm, referred as circadian rhythm, is known to exist in our human body and to consist of two parts. The central clock is located in the suprachiasmatic nucleus in the hypothalamus. There also exist peripheral clocks, the biological clocks in each organ including cardiovascular tissues or cells. The medical literature finds that it is essential for cardiovascular health and disease prevention that all these clocks are tightly synchronized. Daylight is known to play a key role as a zeitgeber (timekeeper) that can exactly synchronize or otherwise reset the central clock (see the literature summarized in Berson et al. (2002)).^2^ Evidence shows that misalignment between the internal clock and external environment, such as light, can induce disorders in cardiovascular organ (Smolensky et al. 2007; Takeda and Maemura 2011; Durgan and Young 2012; Portaluppi et al. 2012; Giuntella and Mazzonna 2019).

Second, DST, directly and indirectly through disrupted circadian rhythm, can also harm the quality and duration of sleep, and loss of duration and efficiency in sleeping is also known to contribute to cardiovascular risks.^3^ Not only can DST directly lead to insufficient sleep particularly in the timing of the springing forward, but disrupted circadian rhythm due to DST is also an important factor affecting sleep habits (for example, Monk and Folkard 1976; Lahti et al. 2006; Kantermann et al. 2007; Harrison 2013; Tonetti et al. 2013), presenting an indirect channel between DST and deprived sleep. The indirect channel makes cardiovascular risk plausible even after the autumn transition. Using data from the American Time Use Survey, Barnes and Wagner (2009) show that the average sleep duration is 40 minutes shorter on the night of the spring transition, whereas no more sleep is gained at the autumn transition despite an extra hour on the clock.^4^ This is particularly significant for the large number of Americans, and Westerners in general, who suffer from chronic sleep deficiency. Cunningham et al. (2016) show that about one third of American adults sleep less than 6 hours, an hour less than what is recommended by the American Academy of Sleep Medicine and the Sleep Research Society (Watson et al. 2015).

Although these studies illustrate plausible channels through which DST could trigger AMI, the medical literature documenting the direct association between the two is relatively scarce and, when it does exist, has generated little consensus and sometimes contradictory results (Manfredini et al. 2019). Table A1 summarize relevant studies. Among them, the most influential study is Janszky and Ljung (2008) who examine the incidences of AMI in Sweden. They estimate the incidence ratio (IR), as measured by the number of AMI incidences in the first week following the DST shifts, divided by the mean occurrence of AMI in the two weeks before and after the transitions. The technique is essentially similar to estimating differences in means or a regression discontinuity (RD) approach, in which any differences in AMI occurrence between immediately following the DST transitions and those in the control period are attributed to the effect of DST.^5^ In total, they observe 47,812 patients for the period of 1987 to 2006. Their results indicate that AMI incidences increased modestly about 5% after the spring transition, whereas there was no statistically significant effect following the autumn transition. These effects are pronounced more among individuals below 65 years old relative to older cohorts and among women after the spring transition.

Janszky et al. (2012) conduct a follow-up study based on a smaller sample in Sweden between 1995 and 2007 but with better information on age and sex, which allows them to identify risk factors leading to variation in vulnerability to clock adjustments. They find similar effect in terms of the incidence ratio after the spring and autumn transitions, yet they find no statistically significant differences in effect over ages above and below 65 or sex. Culíc (2013) utilizes detailed information obtained from 2,412 patients at University Hospital Centre Split in Croatia to identify potential triggers of AMI. The results indicate that AMI patients increased by about 29% and 44% on the first four working days after the spring and autumn transitions, respectively. The effects are larger among male after the spring transitions but among female after the autumn transition, and among individuals employed or engaging in physical activities.

Given the variation in effects over different demographics, it is likely that the magnitudes of effects differ across countries. To date, Jiddou et al. (2013) is the only existing study that provides evidence in the United States. Based on 935 AMI patients presenting to the emergency centers at the Royal Oak and Troy campuses of the Beaumont Hospitals in Michigan from October 2006 to April 2012, they find a statistically significant effect (a 17% increase in AMI) only after the spring transition, though the difference in the estimates between the spring and autumn transitions are not statistically significantly different.

Toro et al. (2015) use daily mortality data on AMI in 2007–2012 in Brazilian states that observe DST and document an approximately 8.5% increase in the mortality rates after the spring transition and a comparable increase of 6.5% after the autumn transition, while they do not find any change in other states where DST was not practiced.

Kirchberger et al. (2015) consider 25,499 cases of coronary death and non-fatal AMI among 25–74 aged population in the region of Augsburg, Germany, between 1985 and 2010. The study finds no overall impact on AMI risk after the spring or autumn transitions, whereas the spring transition appears to increase AMI incidence among men.

Sipilä et al. (2016) use nationwide AMI incidence and in-hospital mortality in Finland in 2001–2009 and finds no significant impact after either the spring or autumn transitions.

Jin and Ziebarth (2020) use AMI hospitalization data from Germany and show a decrease in AMI admissions by 3.6% after the autumn transition, while the effects after the spring transition is not reported.

**Figure A1:**
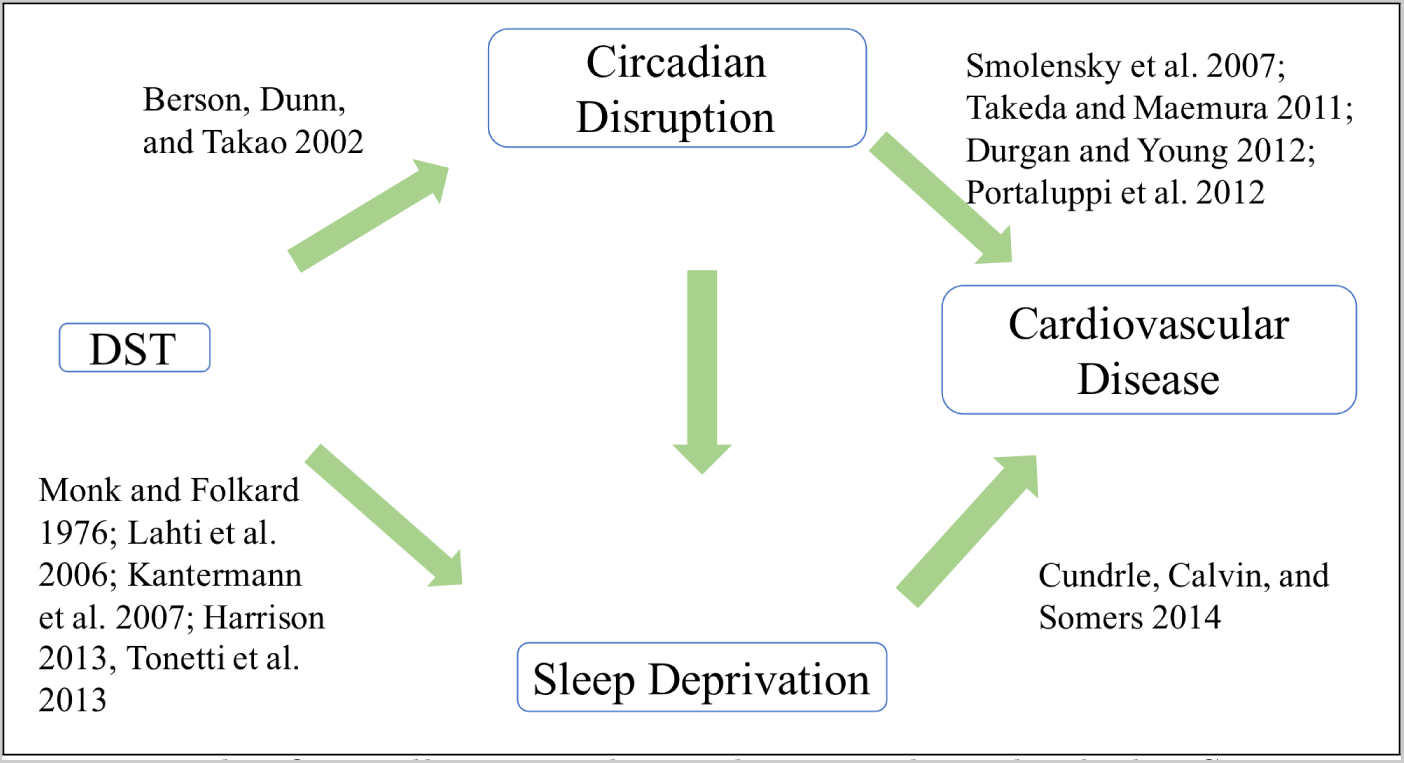
The Linkage between DST and AMI *Notes*: This figure illustrates the mechanisms through which DST can affect AMI from the medical literature.

**Table A1:** Medical Literature on the Linkage between DST and AMI

| Authors | Sample | Results (IR) |  | Risk factors |
| --- | --- | --- | --- | --- |
|  |  | Spring | Fall |  |
| Janszky and Ljung (2008) | 47,812 patients admitted in Sweden in the first week and two weeks before and after the shifts in 1987-2006. | 1.051<br>(1.032–1.071) | 0.98<br>(0.969–1.002) | Age under 65, women for spring and men for autumn |
| Janszky et al. (2012) | 14,521 patients admitted at participating hospitals in Sweden in the first week and two weeks before and after the shifts in 1995-2007. | 1.039<br>(1.003–1.075) | 0.995<br>(0.965–1.026) | No difference over age and gender |
| Čulić (2013) | 2,412 patients hospitalized due to AMI in 1990-1996 at University Hospital Centre Split in Split, Croatia | 1.29<br>(1.09–1.49) | 1.44<br>(1.19–1.69) | Men for spring and women for autumn |
| Jiddou et al. (2013) | 935 patients to the emergency centers at Beaumont Hospitals in Royal Oak and Troy, Michigan. | 1.17<br>(1.00–1.36) | 0.99<br>(0.85–1.16) | N.A. |
| Toro et al. (2015) | 1,995 state-day death cases in Brazil on the day before and after DST in 2007-2012. | 1.085<br>(1.047–1.123) | 1.065<br>(1.006–1.124) | N.A. |
| Kirchberger et al. (2015) | 25,499 coronary deaths and non-fatal AMI cases in the region of Augsburg, Germany in 1985–2010 | 1.077<br>(0.981–1.182) | 1.025<br>(0.928–1.133) | Men |
| Sipilä et al. (2016) | Nationalwide AMI incidence and in-hospital mortality in Finland in 2001–2009. | 0.98<br>(0.94–1.02) | 0.99<br>(0.94–1.04) | No difference over age or gender |
| Jin and Ziebarth (2020) | 336,604 county-day hospitalization cases in Germany in the first week and three weeks before and after DST in 2000-2008. | N.A. | 0.964<br>(0.938–0.990) | N.A. |

*Notes:* This table summarizes the existing studies that examined the effect of DST on AMI. Their results are presented in incidence rates (IR), which compare the incidence rates of events occurring after the DST transition to those in the comparison weeks (typically two or three weeks before and after the transition), thereby the value of greater than one indicates an increased rate, whereas that of lower than one indicates a decreased rate. The respective 95% confidence intervals are presented below the IR. N.A. means not available.

## B. Historical Debates over DST in Indiana

DST was first introduced in Germany in 1916 and was immediately adopted by many other European countries. In the U.S., it was first adopted nationwide in 1918 during World War I for seven months as a means of maximizing use of available sunshine and thereby conserving coal and energy with the slogan “Set the clock ahead one hour and win the war!”^6^ Although DST was immediately repealed by the government after the war due to lack of public support, it was again adopted and continued for the duration of World War II by President Franklin Roosevelt in the form of year-round DST, called “War Time.” The Uniform Time Act of 1966 marked the first attempt by the federal government to establish standard time within time zones and to designate dates on which individual states would be required to turn the clock forward in spring and back in autumn.^7^

Among the historical debates over DST both within and outside the U.S., Indiana presents a unique scenario. Internal disputes over the practice of DST rendered Indiana a special case with multiple time zones and exemption from DST under the Uniform Time Act of 1966.^8^ As a result, 6 northwestern Indiana counties near Chicago and 6 southwestern counties close to Kentucky and southern Illinois adopted Central Standard Time (CST), while the other counties adopted Eastern Standard Time (EST).^9^

The 1972 Uniform Time Act Amendment further permitted multiple time zones and non-uniform practice of DST within states. At that point, 75 Indiana counties in EST decided not to practice DST, while all 12 counties in CST plus 5 counties in EST^10^ started observing DST (Department of Transportation (DOT) 2006). Since then, requests from a few counties to change their time zones have been made several times in Indiana. The only basis on which the DOT was authorized to change time zones was to facilitate or preserve the “convenience of commerce” (DOT 2006). The DOT accepted a petition from Pike County to move from CST to EST in 1977 and Starke in 1991, while it denied requests from 5 southwest counties in 1985 and 2 northwest counties in 1986 to switch to EST. As a result, during the period between 1991 and 2005, 15 out of 92 counties observed DST, while another 77 counties did not.^11^ In 2005, the Indiana state legislature passed a law that required all counties to observe DST starting in 2006. At the same time, 8 counties petitioned the DOT and were approved to move from EST to CST when DST began in April 2006, relieving them of the need to move clocks forward by an hour.^12^ Later in March 2007, Pulaski County changed its time zone back to EST, resulting in a two-hour shift forward, and in November 2007 Daviess, Dubois, Knox, Martin, and Pike counties changed their time zone back to EST, negating the time change when DST ended.

For the purpose of identifying the effect of DST on health, it is worth noting that the series of federal policies regarding DST resulted in splitting Indiana counties into observing and not observing DST before 2006. The variation in time zone and related DST observance status is based on the “convenience of commerce,” as evaluated by the DOT, and thus unrelated to other factors such as health concerns.

**Figure B1:**
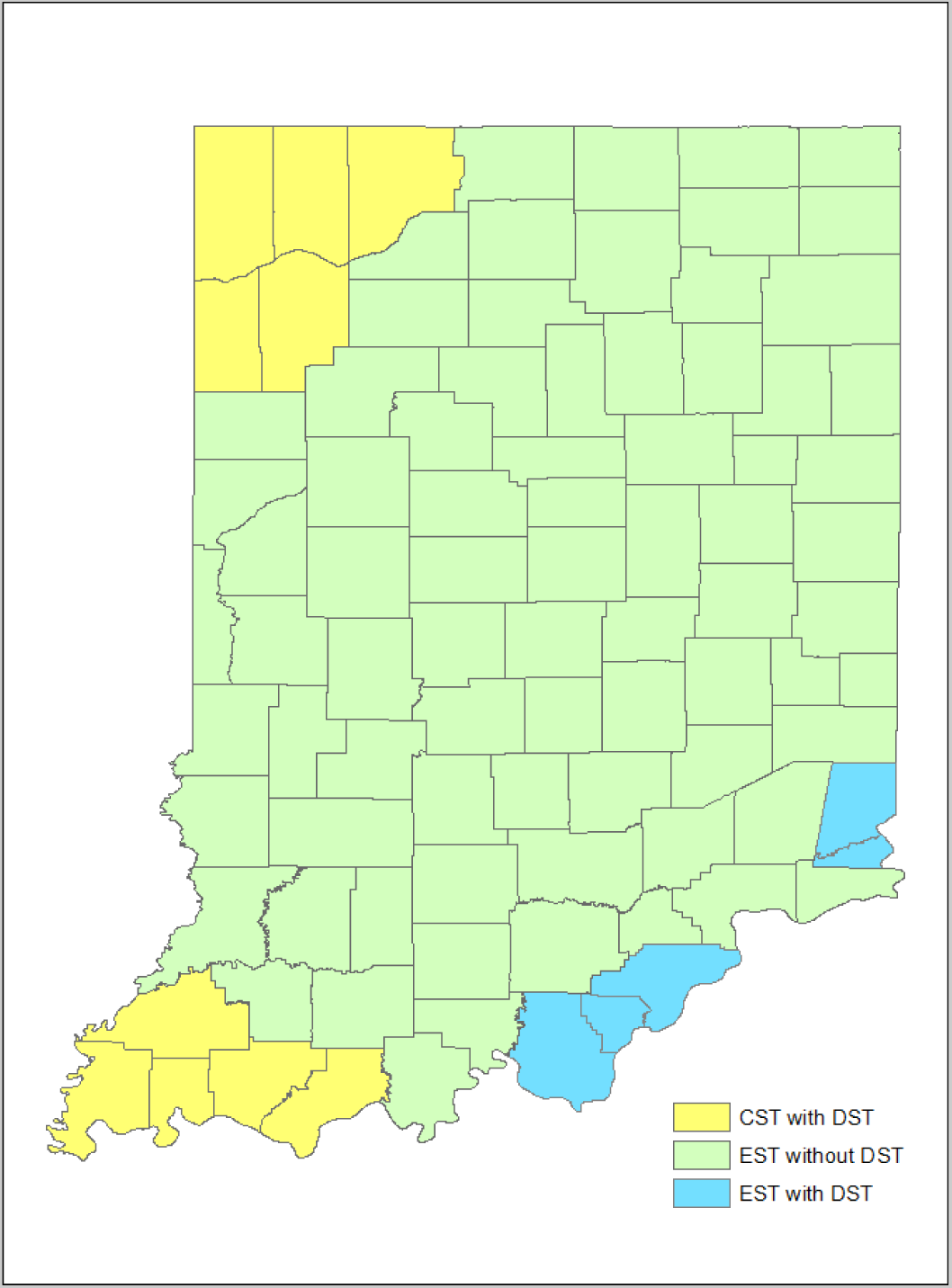
DST Practice and Time Zone in Indiana *Notes*: This map illustrates three types of counties in Indiana as determined by their DST practice and time zone. The counties highlighted by yellow follow the Central Standard Time and practiced DST throughout our study period. The large majority of counties that are highlighted by green follow the Eastern Standard Time and did not practice DST before 2006. The counties with blue follow the Eastern Standard Time and practiced DST throughout our study period.

## C. Additional Information on Data and Summary Statistics

We obtained daily precipitation and temperature information, important factors that change on a daily basis and are likely to be associated with AMI, from NOAA Global Historical Climatology Network Daily (GHCN-D) (NOAA, 2022). The dataset contains daily precipitation and maximum and minimum temperature at the station level. In total, there are more than 1,760 stations across Indiana. We compute the daily average temperature at the station level simply by taking an arithmetic average of maximum and minimum temperature of the day. County level daily temperature and precipitation are then computed by taking an average of all available readings within a county. For dates without any reading in a county, we use the state-wide average information as a proxy in order to preserve the number of observations. There is no day on which none of the stations across the state reports information.

Table C1 describes the mean in the first row, standard deviation in the squared bracket in the second row, and the number of observations in the third row, for the respective variable. The sample includes total cases in the second column, observations 4 weeks before and after the spring transition in the third column, and observations 4 weeks before and after the autumn transition in the fourth column, regardless of whether DST was observed or not. It shows that AMI occurred predominantly to older male individuals: the average age of patients is 53.1 years old, and 71.1% of incidents occurred to men. The average pre-insurance medical cost of treatment is $36,099.9, and the average length of stay for inpatients is 4.1 days. 82.3% of AMI admissions are inpatients. 66.5% of the patients have private insurance, while 13.4% have Medicare, 8.4% have Medicaid, and 7.5% have no insurance. The average number of AMI admissions at the county-daily level is 5.86 per one million population.

**Table C1:** Summary Statistics

| Variable | All | Spring | Autumn |
| --- | --- | --- | --- |
| Age | 53.079<br>[7.858]<br>61,698 | 53.129<br>[7.821]<br>9,747 | 53.283<br>[7.800]<br>9,119 |
| Male (0/1) | 0.711<br>[0.453]<br>61,698 | 0.707<br>[0.455]<br>9,747 | 0.705<br>[0.456]<br>9,119 |
| Total charge (\$) | 36,099.9<br>[31916.4]<br>60,705 | 36,461.3<br>[32008.8]<br>9,603 | 36,613.7<br>[32456.1]<br>8,964 |
| Hospital stay (days) | 4.074<br>[4.084]<br>50,053 | 4.064<br>[4.003]<br>7,955 | 4.223<br>[4.295]<br>7,428 |
| Inpatient (0/1) | 0.823<br>[0.381]<br>61,698 | 0.827<br>[0.378]<br>9,747 | 0.825<br>[0.380]<br>9,119 |
| Medicare (0/1) | 0.134<br>[0.341]<br>61,697 | 0.134<br>[0.341]<br>9,747 | 0.137<br>[0.344]<br>9,119 |
| Medicaid (0/1) | 0.084<br>[0.278]<br>61,697 | 0.085<br>[0.279]<br>9,747 | 0.081<br>[0.272]<br>9,119 |
| Govt. insurance (0/1) | 0.024<br>[0.153]<br>61,697 | 0.023<br>[0.149]<br>9,747 | 0.027<br>[0.161]<br>9,119 |
| Private insurance (0/1) | 0.665<br>[0.472]<br>61,697 | 0.675<br>[0.468]<br>9,747 | 0.659<br>[0.474]<br>9,119 |
| Self pay (0/1) | 0.075<br>[0.263]<br>61,697 | 0.069<br>[0.254]<br>9,747 | 0.077<br>[0.267]<br>9,119 |
| AMI admissions rates | 5.857<br>[15.027]<br>257,232 | 5.880<br>[15.019]<br>40,480 | 5.737<br>[14.768]<br>38,824 |

*Notes:* This table presents the summary statistics for the working-age population between 20 and 64 in three periods: throughout the year in the second column, 4 weeks before and after the spring and autumn DST transitions (regardless of whether it is observed or not) in the third and fourth columns, respectively. For each variable, the first row denotes the mean value, the second row in the square bracket denotes the standard deviation, and the third is the number of observations. The unit of observations is at the patient level, except AMI admission rates, which is at the county-daily level. The AMI admissions rates are per one million working-age population.

## D. Short-term Effects at the Cutoffs

### RD and Additional RDDD results

This section provides additional evidence from the main analysis at the daily level.

Table D1 presents the main RD estimates at the spring transition when DST was practiced in Panel A and the placebo evidence when DST was not practiced in Panel B. Consistent with the graphical evidence in the main text, Panel A documents substantial increases in AMI admissions around the spring transition. The estimate in Column (1) based on Equation (1) suggests that on average the AMI admissions increased by 1.332 cases per one million working-age population when DST starts, which represents an approximately 24.0% increase in the daily AMI admission rates and is statistically significant at the 1 percent level. Column (2) controls for temperature and precipitation, and Column (3) additionally removes cross-county and over-time variation in AMI cases by including the county- and year-fixed effects, but the point estimates are virtually unchanged. Column (4) aggregates the observations to the weekly level in an effort to mitigate day-to-day fluctuations in AMI incidence.The estimated effect is the same as that inferred by the daily analysis, indicating an approximate 16.4% increase in weekly admission rates of AMI. We also explore the effects on AMI severity using total medical charges in Column (5) and length of hospital stay for inpatients in Column (6) at the patient level. We find that the total charges increases by about 10.1%, whereas the length of hospital stay is unchanged.

Panel B illustrates evidence from placebo tests by exploring discontinuities around the DST transition when DST was not practiced. The evidence supports the identification assumption that the observed increase in AMI incidences is absent when DST was not practiced. The point estimates are substantially lower in magnitude, all negative in sign, and not statistically significantly different from zero. Moreover, variables that proxy the severity of AMI cases show no significant discontinuity at the transition either.

Table D2 provides analogous evidence for the autumn transition. We find that the increase in AMI incidence is substantially lower at the autumn transition than at the spring transition. The point estimate in Column (1) suggests that the AMI admissions increased by 0.499 per 1 million population, which corresponds to an approximate 9.25% increase from the average. In contrast, the effect on total charges is larger for the autumn transition than the spring transition. The point estimate suggests that the total charges increased by approximately 10.8%, and is statistically significant at the 10% level. The effect on the length of hospital stay for inpatients remains negligible.^13^

As Figure 1 has already illustrated, Panel B also confirms that the AMI admissions increased by about 0.829 cases per 1 million population when DST ended, even when DST was not practiced. The estimated coefficients in Panel B are consistently greater than those in Panel A across various specifications, suggesting that AMI admissions fell rather than increased due to DST relative to what would have happened had DST not been practiced. In contrast, Column (5) suggests that AMI severity did not increase. If anything, individual cases became less severe around the end of DST.

Figure D1 presents the analogous figure to Figure 1 in the main text but for the total charges.

Figure D2 presents the analogous figure to Figure 1 in the main text but for the length of hospital stay.

Figure D3 presents the analogous figure to Figure 1 in the main text but including weekends in the observations.

Table D3 presents additional results using the RDDD analysis on other dependent variables. In particular, we consider total medical charges and the length of hospital stay as proxies for disease severity, following Dranove et al. (2003). We find suggestive evidence that the total charges increased after the autumn transition, although the point estimate is not statistically significant, whereas they clearly did not change after the spring transition in Column (1). This finding carries two important implications. First, AMI cases induced by the spring transition are no less severe than typical AMI cases arising from other causes. Second, although AMI admission rates did not increase after the autumn transition, the severity of individual cases may have increased. Although the point estimate is not statistically significant, the estimated effect is larger than what is suggested by the simple RD model (which is statistically significant at the 10% level in Online Appendix Table D2) and economically significant at an approximate 17.0% (95%CI [-11.7% 45.7%], *p* = 0.242, *n* = 8,854) increase. The finding suggests that the number of AMI admissions alone may not sufficiently capture the overall impacts of DST on health. We do not find any impacts on length of hospital stay at either spring or autumn transitions in Column (2), suggesting that such increased charges do not reflect longer hospital stays.

**Figure D1:**
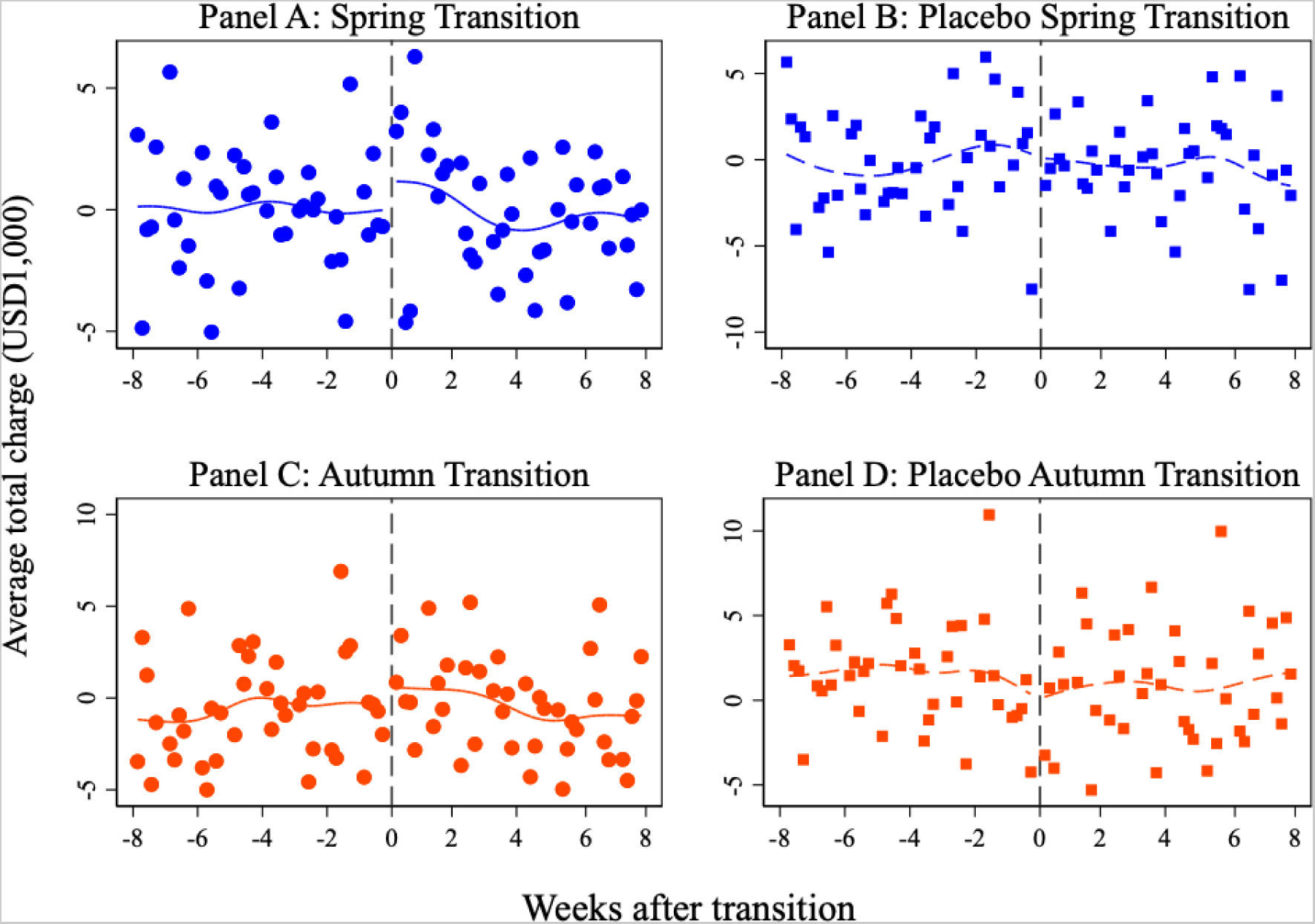
Total Charges around DST *Notes*: This figure plots the daily average of (log) total charges among the working population patients aged between 20 and 64, residualized by the day-of-week effects, county fixed effects, year fixed effects, and temperature and precipitation effects. The level of observations is at the patient level. The lines indicate smoothed local polynomial trends with the bandwidth of seven days. Panels A and C include observations when the respective DST was practiced, whereas Panels B and D include observations when the respective DST was not practiced.

**Figure D2:**
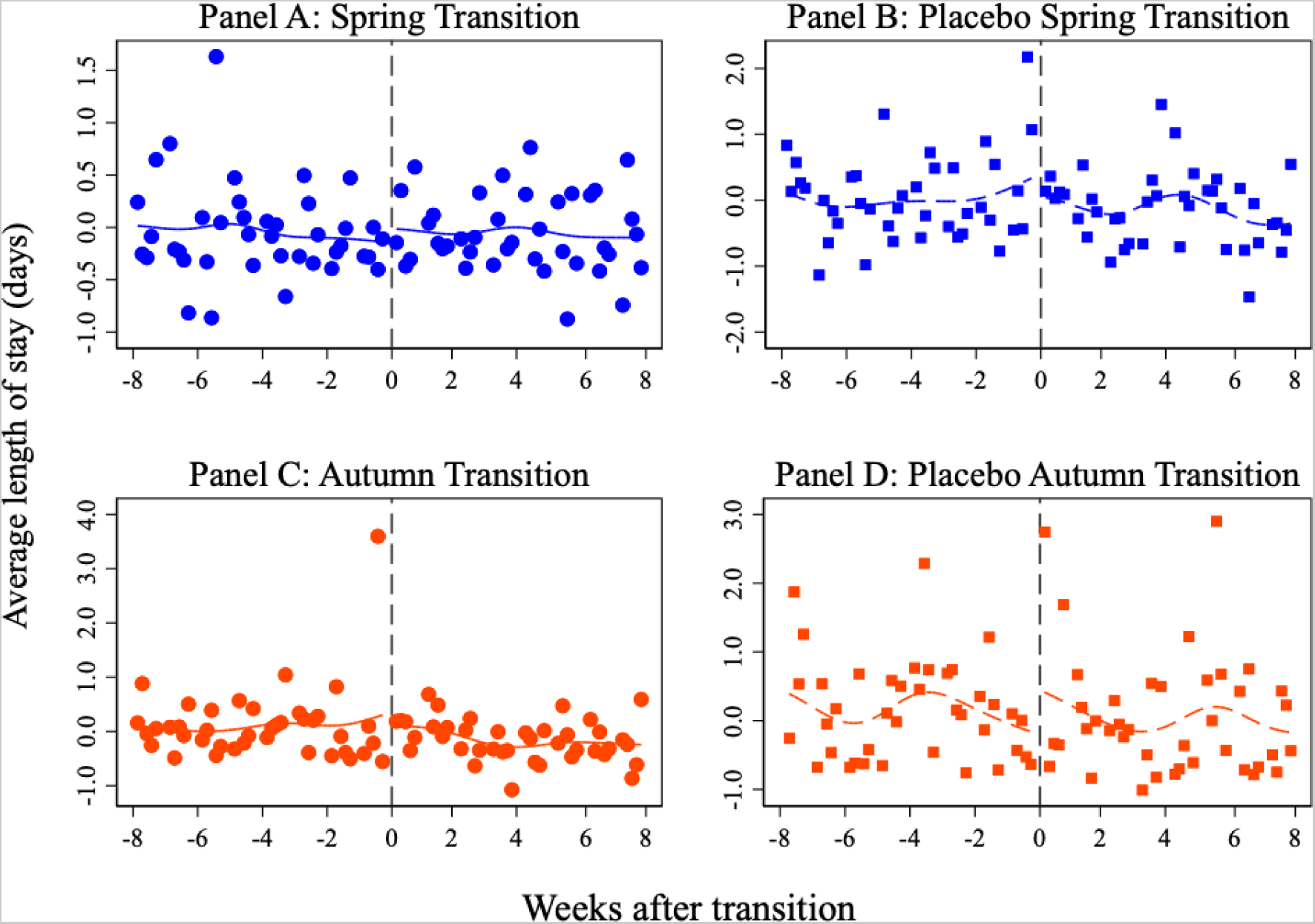
Length of Stay around DST *Notes*: This figure plots the daily average of (log) total length of hospital stay among the working population inpatients aged between 20 and 64, residualized by the day-of-week effects, county fixed effects, year fixed effects, and temperature and precipitation effects. The level of observations is at the patient level. The lines indicate smoothed local polynomial trends with the bandwidth of seven days. Panels A and C include observations when the respective DST was practiced, whereas Panels B and D include observations when the respective DST was not practiced.

**Figure D3:**
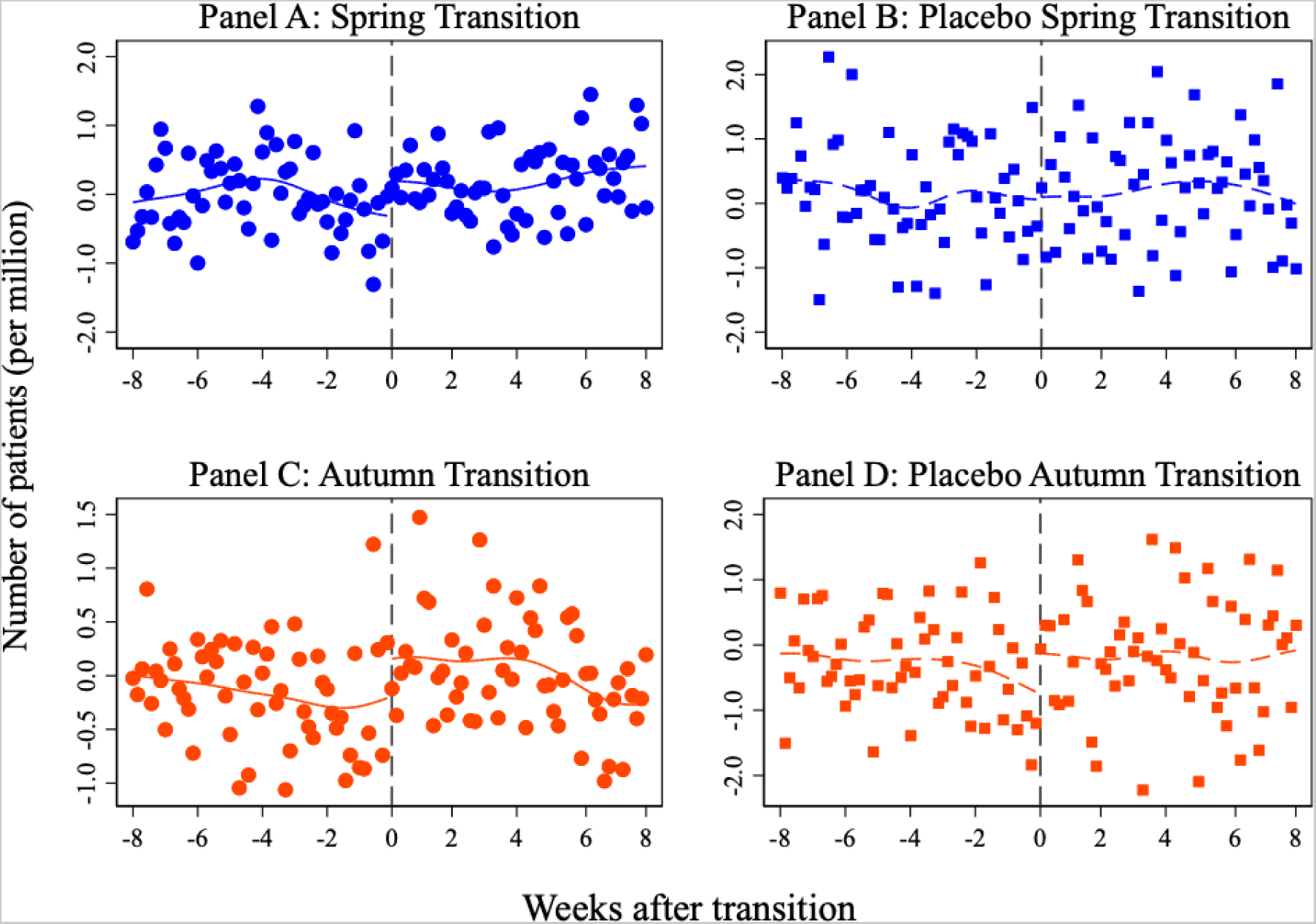
AMI Admission Rates around DST with weekends *Notes*: These figures are analogous to Figure 1 in the main text but including weekends in the observations. The outcome is the daily average AMI admission rates per one million working-age population among the working population aged between 20 and 64, weighted by the working population at the county level, residualized by the day-of-week effects, county fixed effects, year fixed effects, and temperature and precipitation effects. The lines indicate smoothed local polynomial trends with the bandwidth of seven days. Panels A and C include observations when the respective DST was practiced, whereas Panels B and D include observations when the respective DST was not practiced.

**Table D1:** The RD Estimates of the Effects of *Spring* Transition on AMI

|  | Admission rates |  |  |  | Total charges | Length of stay |
| --- | --- | --- | --- | --- | --- | --- |
|  | (1) | (2) | (3) | (4) | (5) | (6) |
| *Panel A: Main* |  |  |  |  |  |  |
| PostTrans | 1.332***<br>(0.343) | 1.142***<br>(0.344) | 1.152***<br>(0.343) | 6.307***<br>(2.093) | 0.101**<br>(0.050) | 0.032<br>(0.033) |
| Mean | 5.548 | 5.548 | 5.548 | 38.467 | 40,416.1 | 4.040 |
| N | 27,840 | 27,840 | 27,840 | 5,568 | 6,603 | 5,322 |
| *Panel B: Placebo* |  |  |  |  |  |  |
| PostTrans | -0.155<br>(0.508) | -0.352<br>(0.510) | -0.355<br>(0.510) | -0.769<br>(2.975) | 0.089<br>(0.095) | -0.061<br>(0.061) |
| Mean | 6.392 | 6.392 | 6.392 | 43.331 | 28,646.7 | 4.545 |
| N | 12,640 | 12,640 | 12,640 | 2,528 | 2,946 | 2,426 |
| Fixed effects | N | N | Y | Y | Y | Y |
| Controls | N | Y | Y | Y | Y | Y |
| Level of obs. | Daily | Daily | Daily | Weekly | Individual | Individual |

**Table D2:** The RD Estimates of the Effects of *Autumn* Transition on AMI

|  | Admission rates |  |  |  | Total charges | Length of stay |
| --- | --- | --- | --- | --- | --- | --- |
|  | (1) | (2) | (3) | (4) | (5) | (6) |
| *Panel A: Main* |  |  |  |  |  |  |
| PostTrans | 0.499* | 0.300 | 0.296 | 3.259* | 0.108* | 0.032 |
|  | (0.292) | (0.292) | (0.292) | (1.824) | (0.061) | (0.047) |
| Mean | 5.393 | 5.393 | 5.393 | 36.811 | 39,982.3 | 4.285 |
| N | 26,622 | 26,622 | 26,622 | 5,592 | 6,169 | 5,052 |
| *Panel B: Placebo* |  |  |  |  |  |  |
| PostTrans | 0.967* | 0.812 | 0.829 | 5.155* | -0.062 | 0.039 |
|  | (0.525) | (0.526) | (0.526) | (2.684) | (0.115) | (0.058) |
| Mean | 6.072 | 6.072 | 6.072 | 39.558 | 29,256.3 | 4.643 |
| N | 12,202 | 12,202 | 12,202 | 2,504 | 2,685 | 2,210 |
| Fixed effects | N | N | Y | Y | Y | Y |
| Controls | N | Y | Y | Y | Y | Y |
| Level of obs. | Daily | Daily | Daily | Weekly | Individual | Individual |

**Table D3:**
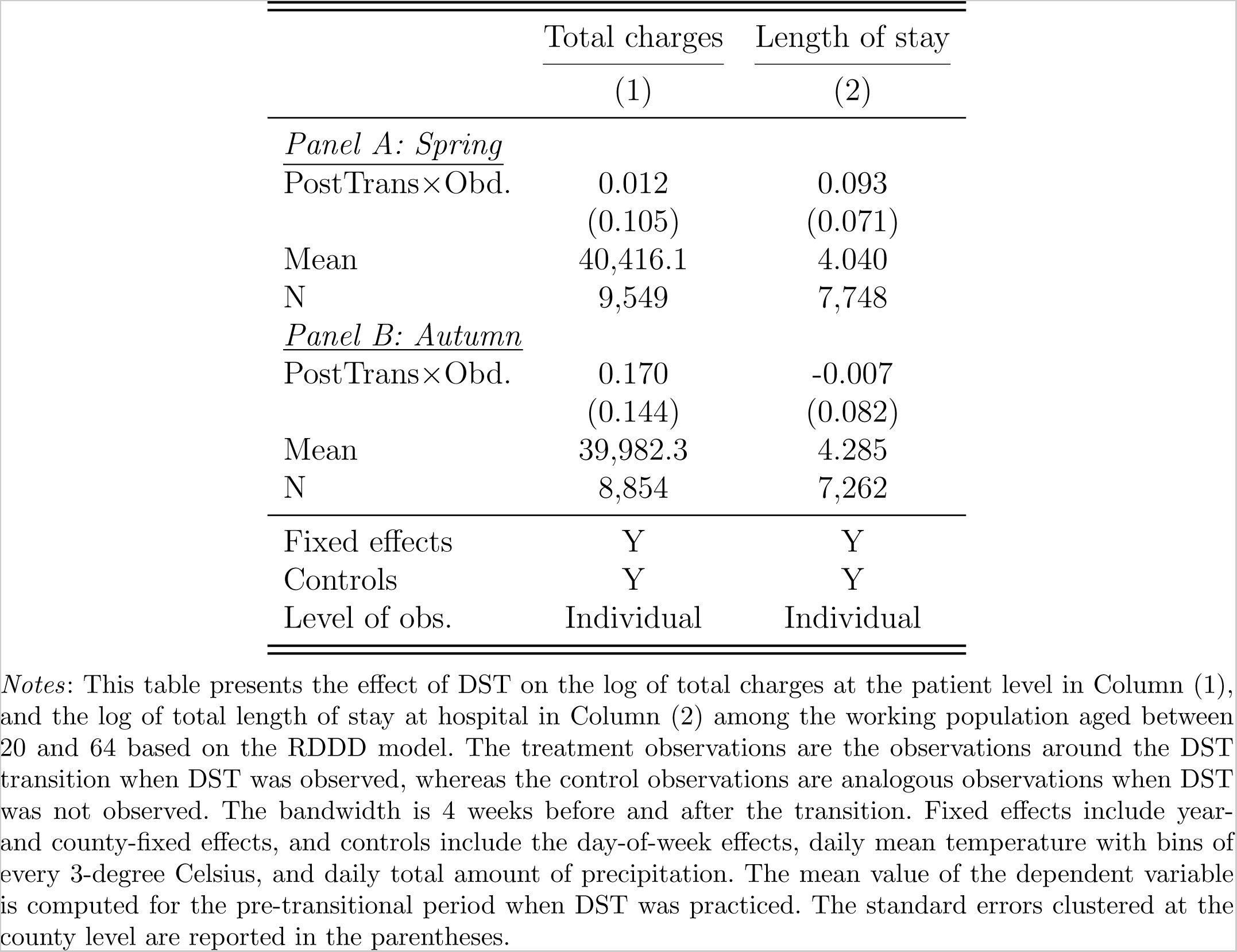
The RDDD Estimates of the Effect of DST Transition on AMI

### Weekly results

This subsection provides additional evidence from the main analysis at the weekly level. The observations on the number of admissions are aggregated to the weekly level to reduce the day-to-day fluctuations in the number of AMI admissions. Here we consider only AMI admissions, as it makes no sense to aggregate patient-level total charges or length of hospital stay to the weekly level. The number of weeks since the DST transition is used as the running variable. The sample does not include weekends or holidays to be consistent with the main analysis. The results are similar to the daily-level of analysis.

**Figure D4:**
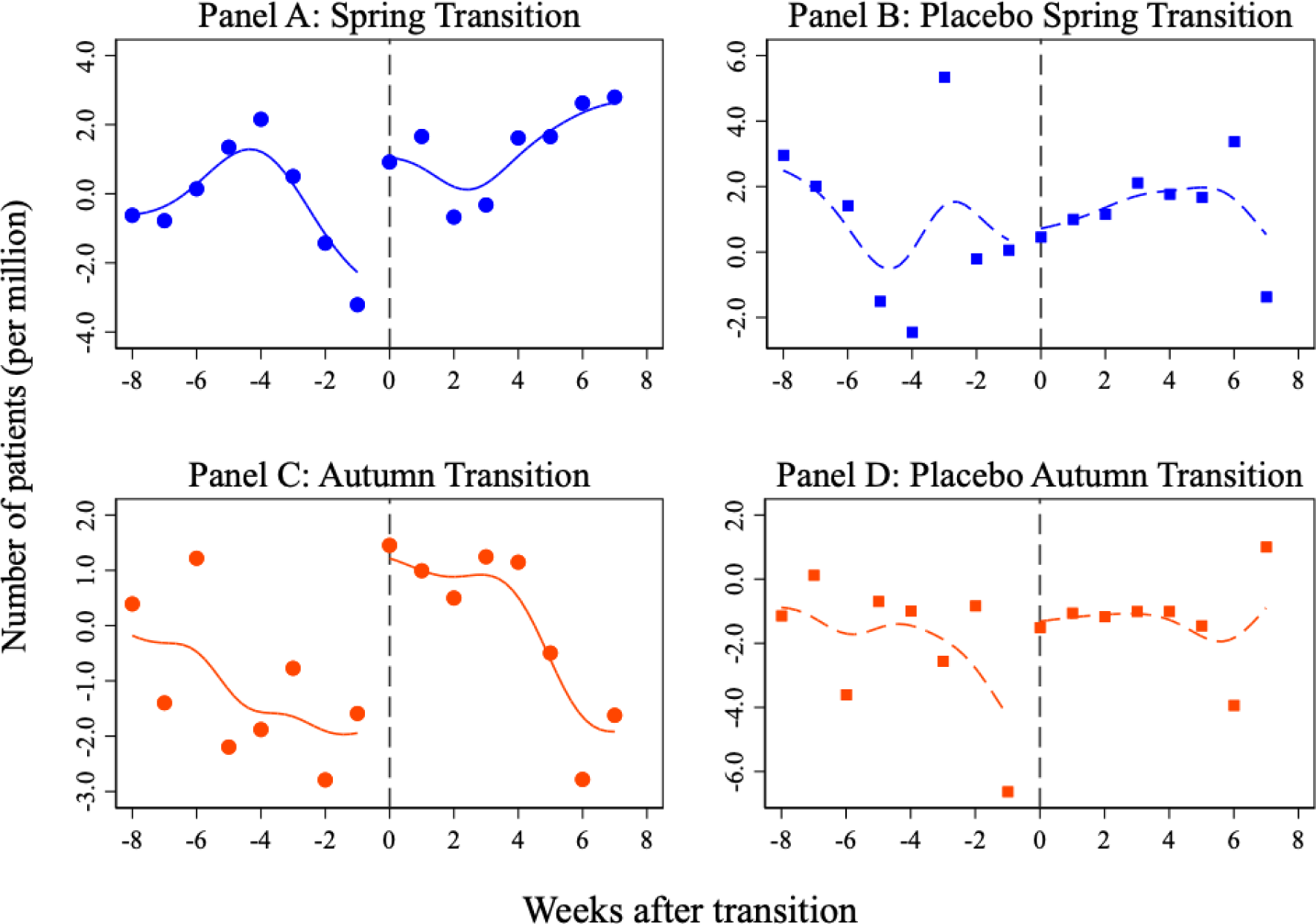
Weekly AMI Admissions around DST *Notes*: This figure plots the weekly average admission rates per one million working-age population among the working population aged between 20 and 64, residualized by the day-of-week effects, county fixed effects, year fixed effects, and temperature and precipitation effects, and weighted by the working-age population at the county level. The level of observations is at the county-weekly level. The lines indicate smoothed local polynomial trends with the bandwidth of seven days. Panels A and C include observations when the respective DST was practiced, whereas Panels B and D include observations when the respective DST was not practiced.

**Table D4:** The RD Estimates of the Effects of *Spring* Transition on *Weekly* AMI Admissions

|  | Admission rates |  |  |
| --- | --- | --- | --- |
|  | (1) | (2) | (3) |
| *Panel A: Main* |  |  |  |
| PostTrans | 6.173***<br>(2.076) | 6.429***<br>(2.084) | 6.307***<br>(2.093) |
| Mean | 38.467 | 38.467 | 38.467 |
| N | 5,568 | 5,568 | 5,568 |
| *Panel B: Placebo* |  |  |  |
| PostTrans | -0.665<br>(2.944) | -0.648<br>(2.978) | -0.769<br>(2.975) |
| Mean | 43.331 | 43.331 | 43.331 |
| N | 2,528 | 2,528 | 2,528 |
| Fixed effects | N | N | Y |
| Controls | N | Y | Y |
| Level of obs. | Weekly | Weekly | Weekly |

**Table D5:** The RD Estimates of the Effects of *Autumn* Transition on *Weekly* AMI Admissions

|  | Admission rates |  |  |
| --- | --- | --- | --- |
|  | (1) | (2) | (3) |
| *Panel A: Main* |  |  |  |
| PostTrans | 3.282* | 3.322* | 3.259* |
|  | (1.827) | (1.828) | (1.824) |
| Mean | 36.811 | 36.811 | 36.811 |
| N | 5,592 | 5,592 | 5,592 |
| *Panel B: Placebo* |  |  |  |
| PostTrans | 4.761* | 5.145* | 5.155* |
|  | (2.675) | (2.686) | (2.684) |
| Mean | 39.558 | 39.558 | 39.558 |
| N | 2,504 | 2,504 | 2,504 |
| Fixed effects | N | N | Y |
| Controls | N | Y | Y |
| Level of obs. | Weekly | Weekly | Weekly |

**Table D6:** The RDDD Estimates of the Effects of DST Transition on *Weekly* AMI Admissions

|  | (1) | (2) | (3) |
| --- | --- | --- | --- |
| <u>Panel A: Spring</u> |  |  |  |
| PostTrans×Obd. | 6.837* | 7.077** | 7.076** |
|  | (3.444) | (3.460) | (3.453) |
| <u>Panel B: Autumn</u> |  |  |  |
| PostTrans×Obd. | -1.479 | -1.823 | -1.896 |
|  | (3.217) | (3.224) | (3.219) |
| Fixed effects | N | N | Y |
| Controls | N | Y | Y |
| Level of obs. | Weekly | Weekly | Weekly |

*Notes:* This table presents the effects of DST on the weekly admission rates per one million working-age population at the county level. The bandwidth is 4 weeks. The regressions are weighted by the working population at the county level. Fixed effects include year- and county-fixed effects, and controls include the day-of-week effects, daily temperature with bins of every 3-degree Celsius, and total amount of precipitation. The standard errors clustered at the county level are reported in the parentheses.
\*\*\* $p < 0.01$ \*\* $p < 0.05$ \* $p < 0.10$

### Robustness

In this section, we attest the robustness of the RDDD results in the main text to various other specifications. All results are presented in Table D7, except for alternative bandwidths whose results are in Table D8.

#### Count model

We use the count variable, e.g., the number of admissions, instead of admission rates for the measure of AMI admissions. Poisson and negative binomial regressions are preferred to OLS to model the count data. We first present the results using the poisson specifications with normalization by including the number of the working-age people as an exposure variable so as to arrive at population-normalized counts (with its logged variable being equal to one). The estimate in the first row based on the poisson regression indicates statistically significant increases in AMI admissions at the 1% level. Poisson regressions perform well conditional on equality of the mean and variance. In our data, overdispersion tests reject equidispersion with a dispersion parameter of approximately 0.53. In this case, poisson regressions lead to a greater Type I error than should occur, meaning the null hypothesis would be more likely to be rejected when it is true. Thus, we employ the negative binomial specifications that perform better when the data are overdispersed. The estimate based on negative binomial regressions in the second row with normalization suggests that AMI admissions increased by about 30.9% and are statistically significant at the 1% level.^14^ The magnitude of the effect is larger yet quantitatively similar to what is suggested by OLS regressions in the main text.

#### Alternative bandwidths

We conduct the main analysis using various other bandwidths: those selected by Ludwig and Miller (2007), Imbens and Kalyanaraman (2012), MSE-optimal bandwidth selector (Calonico et al. 2014), a half size of MSE, twice larger than MSE, 2 weeks, and 6 weeks. Since these bandwidth selectors are adapted to the RD analysis rather than RDDD, we present both results using the same bandwidths selected from the RD analysis in Table D8. All estimated effects remain in the quantitatively similar range of the main analysis and, in most cases, are statistically significant.

#### Including weekends

We include weekends and holidays in the analysis. The average AMI rate on the first day of spring transition appears to be the same as the day before, suggesting that the effect of DST kicks in from the subsequent Monday (Figure D3). Accordingly, the point estimate is lower, yet the magnitude remains in the range of the main estimate and statistically significant at the 5% level in row (3) of Table D7. The evidence that AMI admissions are less likely to be induced by DST on weekends indicates that sleep deprivation appears to be an important mechanism triggering AMI. Indeed, the average AMI rates continue to be lower on weekends than weekdays, suggesting that the effects of sleep deprivation in the first week entering DST does not extend to the following weekends. The evidence suggests that sleep deprivation on the day or its accumulation over a few days on weekdays is likely to induce AMI, whereas extra time to sleep on weekends is likely to mitigate the effect.

#### Polynomial running variable

The local linear regressions perform better in the RD contexts than higher order polynomials (Gelman and Imbens 2019). Nonetheless, we use a third order polynomial to allow non-linear trends in the running variable. The point estimate in row (4) is amplified, although it is less precisely estimated.

#### Alternative control groups

With the inclusion of the county-fixed effects, the main RDDD analysis exploits variation in the DST observation status over time within the county, which essentially utilizes observations in their own counties when DST was not practiced as a counterfactual. With the inclusion of year-fixed effects, which capture the overall time trend, this assumption seems valid. We still experiment with an alternative construction of the control observations. In particular, using observations only before the policy change in 2006, we conduct the RDDD analysis without including the county-fixed effects yet with year-fixed effects. This effectively examines the effect of DST in the counties that had already practiced DST before the policy change by comparing them with the counties that did not practice DST in the respective year. An additional benefit in this analysis is that exactly the same dates are used to define the beginning of the spring transition in each year. The estimated effects in row (5) remain similar in magnitude.

#### Commuting effect

It is likely that counties that did not practice DST before the policy change and are adjacent to counties that did practice DST may have been already effectively affected by DST if people there worked in counties with DST. Thus, we now conduct the analysis only among the counties that did not practice DST before 2006, and separately for counties that are and are not adjacent to counties that practiced DST throughout (see their locations in Figure D5). With commuting effects present, the main analysis understates the true DST effect by differencing out the DST spillover effects. Thus, we expect the effects to be lower among the adjacent counties than non-adjacent counties. The estimated effects are larger for adjacent counties, indicating that such commuting effects are negligible.

**Figure D5:**
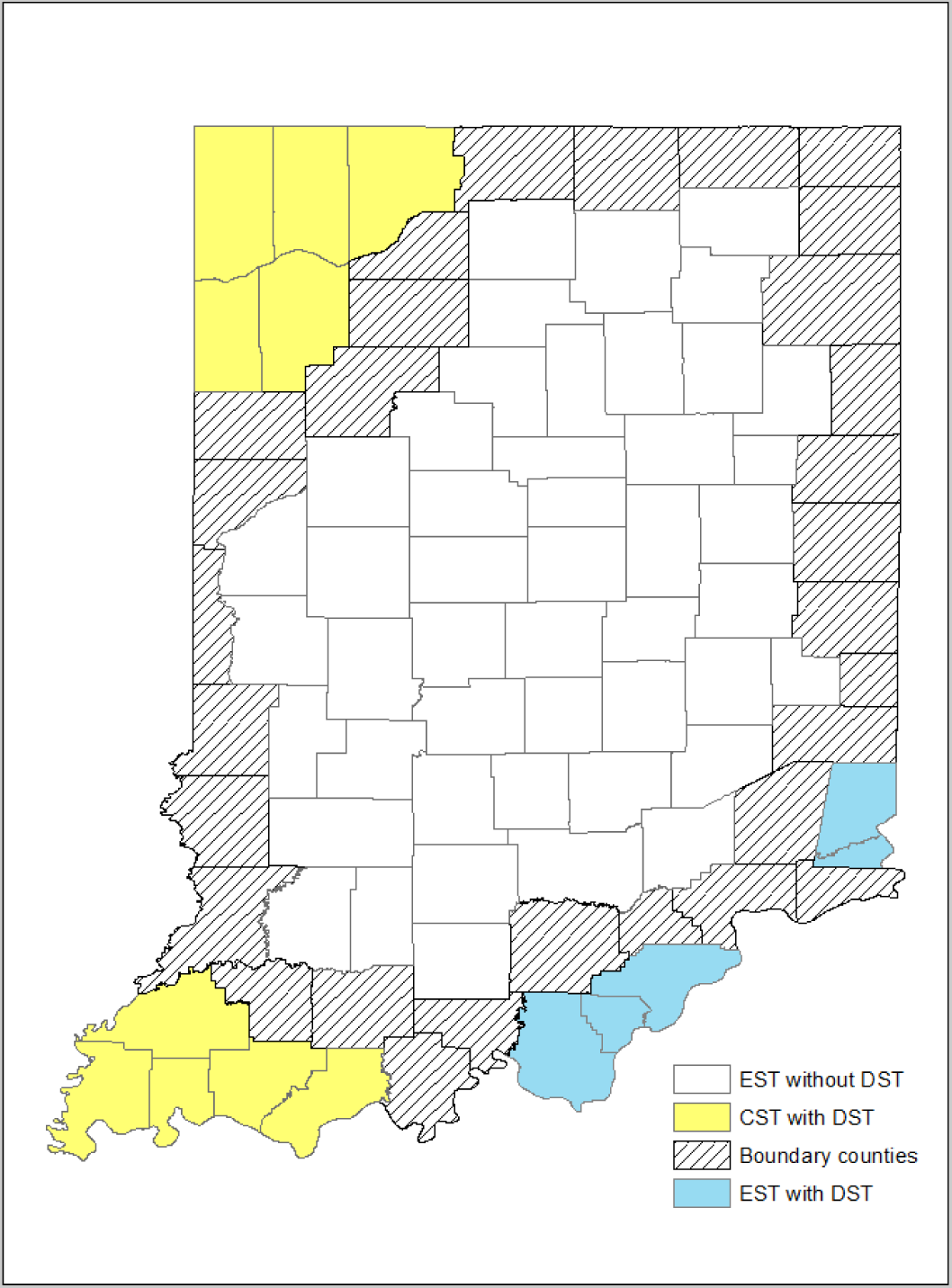
The Location of Adjacent Counties *Notes*: This figure shows the locations of counties that are and are not adjacent to counties that have practiced DST over the study period.

**Table D7:** Robustness: The Effect of Spring Transition on AMI Admissions

|  | Specification | Estimates |
| --- | --- | --- |
| (1) | Poisson | 0.199***<br>(0.061) |
| (2) | Neg. bin. | 0.220***<br>(0.059) |
| (3) | With weekends | 1.056**<br>(0.512) |
| (4) | 3rd order polynomial | 2.756*<br>(1.490) |
| (5) | Alternative control | 1.267<br>(0.852) |
| (6) | Not adjacent | 1.231<br>(0.746) |
| (7) | Adjacent | 2.142**<br>(1.016) |

*Notes:* This table presents the RDD estimates of the effects on AMI admission rates at the spring transition using various alternative specifications. The specifications are from the top row; (1) Poisson regressions are used for the count variable of the number of admissions; (2) Negative binomial regressions are used for the count variable of the number of admissions; (3) The sample including weekends and holidays is used; (4) 3rd order polynomial function in the running variable is used; (5) The sample before 2006 is used without the county fixed effects; (6) Counties that did not practice DST before 2006 and not adjacent to counties that practiced DST throughout the study period are used; (7) Counties that did not practice DST before 2006 and adjacent to counties that practiced DST throughout the study period are used.
\*\*\* $p < 0.01$ \*\* $p < 0.05$ \* $p < 0.10$

**Table D8:** The Estimates based on Alternative Bandwidths

|  | (1) | (2) | (3) | (4) | (5) | (6) | (7) |
| --- | --- | --- | --- | --- | --- | --- | --- |
| *Panel A: RD* |  |  |  |  |  |  |  |
| PostTrans | 1.163***<br>(0.354) | 1.098***<br>(0.397) | 1.227***<br>(0.391) | 1.220**<br>(0.509) | 0.891**<br>(0.348) | 0.877**<br>(0.394) | 0.789**<br>(0.349) |
| N | 28536 | 22968 | 18792 | 9048 | 35496 | 13920 | 41760 |
| *Panel B: RDDD* |  |  |  |  |  |  |  |
| PostTrans×Obd. | 1.353**<br>(0.522) | 1.268**<br>(0.625) | 0.966<br>(0.745) | 2.057*<br>(1.234) | 1.142**<br>(0.484) | 1.340<br>(0.884) | 0.819*<br>(0.464) |
| N | 41492 | 33396 | 27324 | 13156 | 51612 | 20240 | 60720 |
| Selector | LM | IK | MSE | MSE * 1/2 | MSE * 2 |  |  |
| Bandwidth | 29 | 23 | 18.23 | 9.11 | 36.46 | 2 weeks | 6 weeks |

*Notes:* This table presents the estimated effects of the Spring transition based on the RD analysis in Panel A and the RDDD analysis in Panel B using alternative bandwidths. The outcome variable is AMI admission rates per one million working-age population. The level of observation is the daily-county. Column (1) uses the bandwidth selected by Ludwig and Miller (2007), Column (2) by Imbens and Kalyanaraman (2012), Column (3) uses MSE-optimum bandwidth selector (Calonico et al. 2014), Column (4) uses a half size of Column (3), Column (5) uses twice larger than Column (3), Column (6) uses 2 weeks, and Column (7) uses 6 weeks.
\*\*\* $p < 0.01$ \*\* $p < 0.05$ \* $p < 0.10$

### Permutation Test

The key identification assumption for the findings in the main text to offer a causal inference is that observations when DST was not practiced adequately capture the unobserved seasonality effect when DST was practiced. To test the validity of the identification assumption and to put the estimated effects into context, we conduct the permutation test. The basic idea is to assign the placebo DST transition on each date (e.g., 40th day from the actual DST transition as the placebo cutoff) and conduct the same RDDD analysis based on Equation (4) in the main text, and in doing so, we omit days immediately after the actual DST dates to avoid capturing the true DST effect. Following Smith (2016), we exclude the first two weeks after the DST transition as the immediate neighborhood that may be influenced by the true DST effect (e.g., the first two weeks after the spring transition for the effect on AMI admissions or the first two weeks after the autumn transition for the effect on total charges) or any dates that include the 4-week bandwidth in this period (e.g., from 338 days after the actual DST transition to 41 days after the actual DST transition).

Figure D6 plots the distribution of estimated coefficients from the permutation test on the AMI admissions. The vertical dash line indicates the DST effect estimated from the RDDD analysis using the actual DST spring transition. Clearly, the estimated discontinuity at the true transition is extremely large relative to other effects estimated at placebo cutoffs with the implied *p*-value of 0.0068. The evidence that such observed effect at the true DST transition is a rare event also provides support to the identification assumption that the trends in AMI admissions are similar on other dates in this time frame between the observations with and without DST practice.

**Figure D6:**
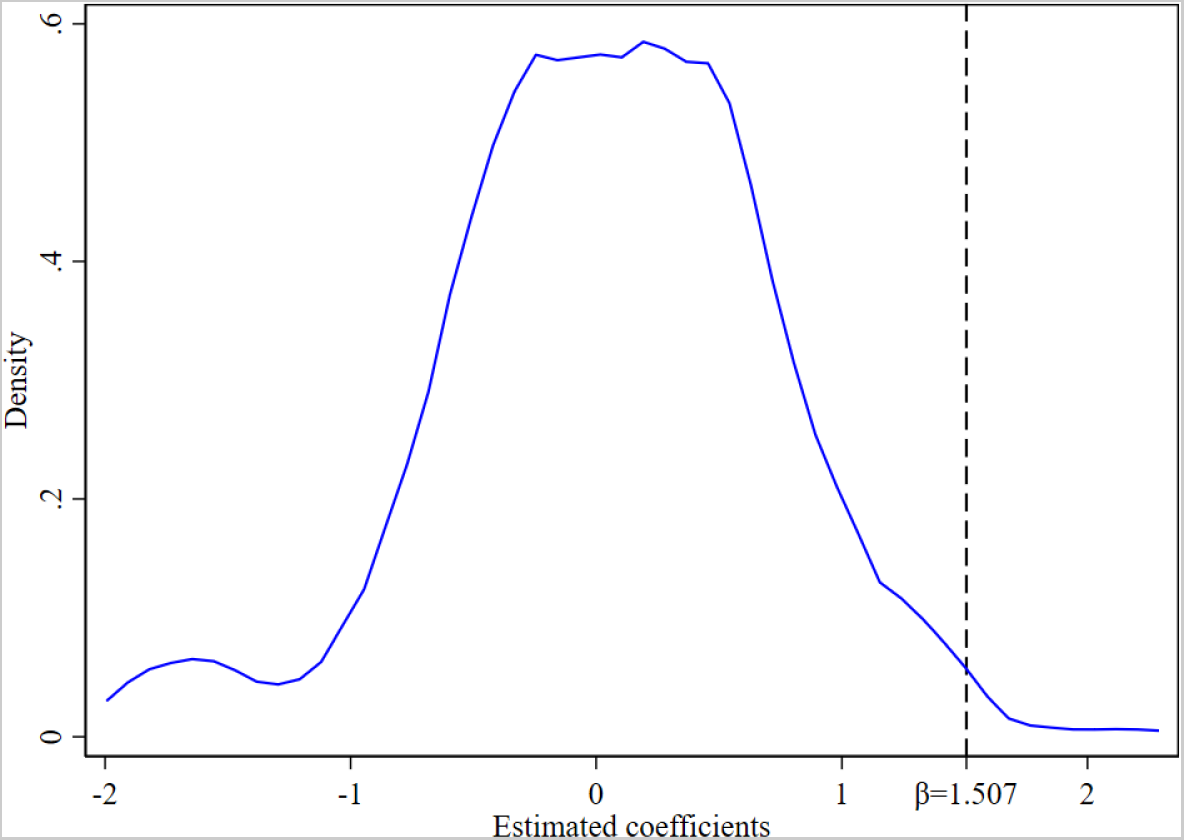
Permutation Test: Distribution of Coefficients on Number of AMI Admissions *Notes*: This figure plots the distribution of coefficients estimated from the permutation test on AMI admission rates per one million working-age population that assigns the placebo DST date to 296 days (from 42 days to 337 days after the true DST transition) outside the 2-week period following the spring transition or any dates that include the 4-week bandwidth in this period. The kernel density is estimated by using Epanechnikov kernel function. The vertical dash line indicates the estimated effect in the main RDDD analysis. The implied *p*-value from this permutation test is 0.0068.

### Heterogeneous Effects of DST

This subsection explores potential heterogeneities in the effects of DST on AMI across various dimensions by applying the main RDDD analysis to a set of subsamples. We examine here only the effects on the number of admissions after the spring transition based on the main results.

On the top row of Figure D7 is the estimate from the main analysis for the purpose of reference. In the second and third rows, we disaggregate the sample into inpatients and outpatients and find that the main effect is mostly driven by the increase in the number of inpatients rather than of outpatients. This is consistent with the fact that a large majority of AMI cases result in hospitalization and with our finding above that AMI cases induced by the spring transition are as severe as typical AMI cases. Still note that the effect on outpatients is not negligible. Although the point estimate is smaller, it indicates an increase of 38.1% from the mean and is statistically significant at the 10% level.

In the following two rows, we explore heterogeneities by gender. While the magnitudes of the estimates are similar between men and women, the impact on men is smaller relative to the mean, and noisily estimated, indicating about an 19.0% increase. On the other hand, AMI admissions increased significantly among women by about 46.5%. The finding that women are more adversely affected by the spring transition than are men is consistent with Janszky and Ljung (2008), although the precise mechanism is unknown. In contrast, Janszky et al. (2012) find no difference by gender, whereas Čulić (2013) finds impacts on men for the spring transition and women for the autumn transition (although not reported, we find no evidence of increased AMI admissions among either men or women at the autumn transition).

In the next two rows below, we compare admissions above and below 50 years old, the approximate mean age of our sample.^15^ We find that the effects are concentrated on people above 50 years old, which is consistent with the fact that AMI is more common among the older population. Although not reported here, we find that DST has no impact on AMI admissions among seniors, as distinct from older working adults, despite the fact that slightly more than a half of AMI cases in Indiana during our study period occurred to seniors above the age of 65 (53.5%). Thus, DST is likely to trigger AMI among the older working population but not among seniors who are likely to be already retired.

Next, we compare patients by insurance type, i.e., between Medicaid and private insurance. The patients on Medicaid experienced a small and insignificant increase in incidences of AMI, whereas patients on private insurance experienced a significant increase in such incidences. Although the precise mechanisms remain unclear, the differential impacts by insurance type may be explained by the fact that people on private insurance are more likely to be full-time workers engaging in jobs with less flexible work schedules that are not aligned with their biological rhythms, while those reliant on Medicaid have more opportunity to adjust to the time shift.

Motivated by the evidence above, we also explore counties by employment rates (e.g., those above and below the mean employment rates) in the bottom two rows.^16^ Although we do not find significant differences here, the point estimate is statistically significant only for counties above the mean employment rate. The estimated effect for counties with lower employment rates is slightly larger but is less precisely estimated. The employment status itself does not indicate the employment sector or whether work is full- or part-time. Unfortunately, we do not have data regarding employment status or work sector in our patient data to allow further investigation.

**Figure D7:**
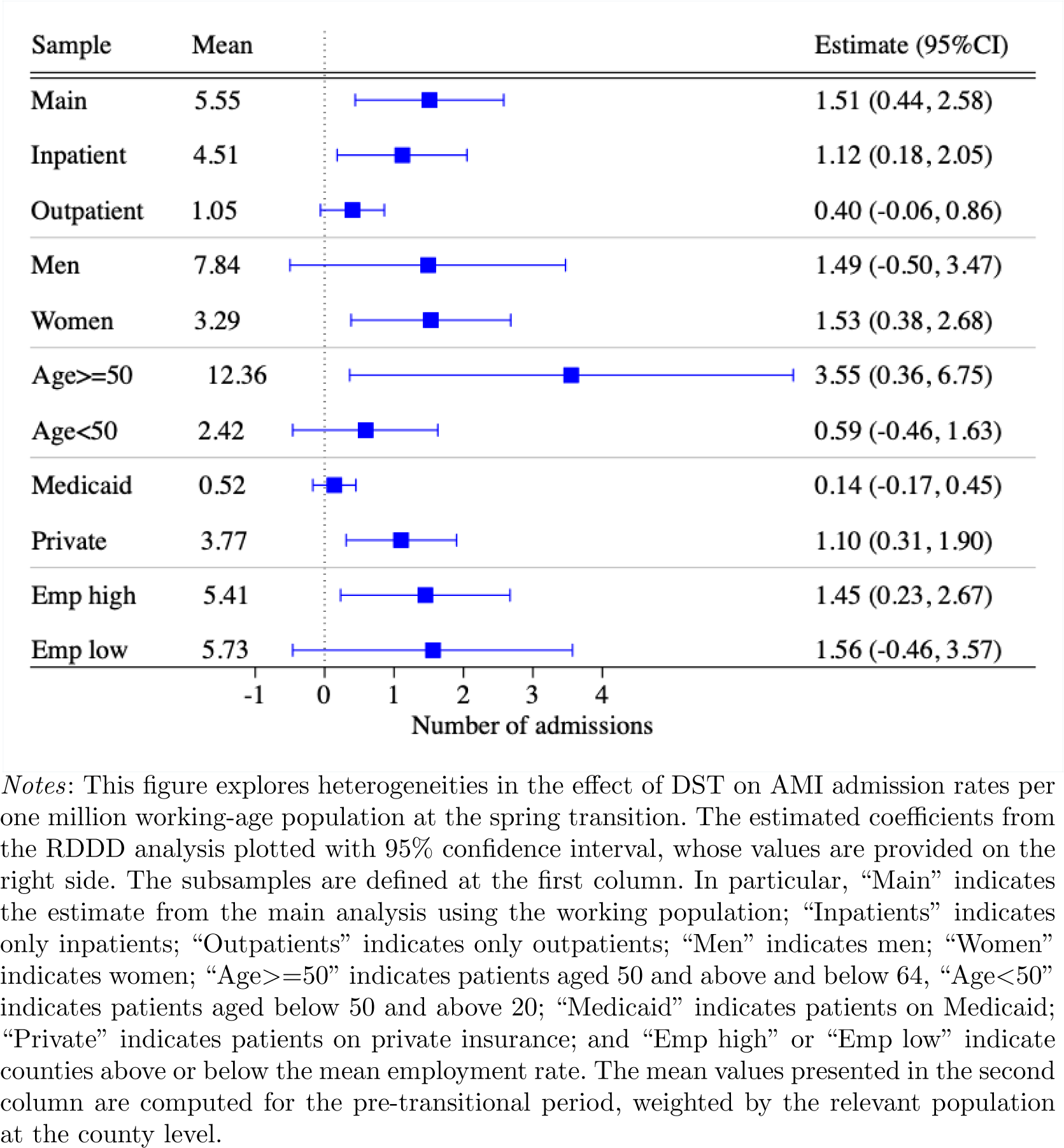
Heterogeneous Effects on Number of Admissions *Notes*: This figure explores heterogeneities in the effect of DST on AMI admission rates per one million working-age population at the spring transition. The estimated coefficients from the RDDD analysis plotted with 95% confidence interval, whose values are provided on the right side. The subsamples are defined at the first column. In particular, “Main” indicates the estimate from the main analysis using the working population; “Inpatients” indicates only inpatients; “Outpatients” indicates only outpatients; “Men” indicates men; “Women” indicates women; “Age*>*=50” indicates patients aged 50 and above and below 64, “Age*<*50” indicates patients aged below 50 and above 20; “Medicaid” indicates patients on Medicaid; “Private” indicates patients on private insurance; and “Emp high” or “Emp low” indicate counties above or below the mean employment rate. The mean values presented in the second column are computed for the pre-transitional period, weighted by the relevant population at the county level.

### Role of Fatal Cases

Our analysis considers only non-fatal infarctions since our data come from hospital discharge and outpatients’ records. In this subsection, we attest that our focus on non-fatal incidences does not undermine the novel conclusions of the work performed.

First, it is not unusual to focus either on non-fatal or on fatal infarctions in the literature of DST and AMI. For example, in the literature summarized in Table A1, Jiddou et al. (2013); Čulić (2013) finds only increased non-fatal admissions but not fatal admissions; Kirchberger et al. (2015) and Sipilä et al. (2016) find no impact on fatal and non-fatal AMi cases; Toro et al. (2015) focuses only on fatal cases; and neither Janszky and Ljung (2008), Janszky et al. (2012), nor Jin and Ziebarth (2020) states whether the hospital admissions involved fatal or non-fatal infarctions. Thus, no study has substantiated that considering fatal cases alters the implications for non-fatal cases.

Second, one may be concerned that fatal cases would truncate the non-fatal observations in our data, which would alter our conclusions. However, our results indeed have implications for fatal cases and suggest that such a concern is not well-founded. Unlike the previous studies on this topic, one of our strengths is to observe hospital medical charges and length of hospital stay as measures of disease severity. Our results show no impacts on either measure in the short-term effect. If the marginal cases who would have barely survived with a heart attack died because the heart attack was intensified by DST, we would expect to see lower disease severity among people who survived and remained in our data. However, our estimated effect on disease severity is virtually zero, whose evidence is consistent with Jiddou et al. (2013), Kirchberger et al. (2015), and Sipilã et al. (2016) who find no impact on fatal incidences.

Third, increased fatal cases, if any, would go against finding increased hospital admissions, thus the only result would be in understating the true effects on the number of hospital admissions. No study has found decreased mortality at the DST transition, and thus lower mortality would not explain increased hospital discharges. Alternatively, DST may hasten and weaken a heart attack that would have resulted in a fatal case several days later, yet not only is there no scientific evidence that hastened heart attack is weakened but also we find no evidence of displacement in hospital admissions, making it unlikely that there is such a displacement effect for mortality.

Fourth, as direct evidence, we have also investigated the effects of DST on AMI mortality in the U.S. The data come from the CDC National Vital Statistics System Multiple Cause of Death (NVSS MCOD) (Centers for Disease Control and Prevention 2011), which reports all AMI fatal cases in 2002–2011 across the U.S. except states that did not practice DST during the study period (without state identities), e.g., Hawaii, Arizona, Indiana, and Alaska.

Because the analysis requires restricted-use data of the exact date of occurrence to construct the daily death counts from AMI, these data were accessed through the Research Data Center.^17^ Using the standard RD specification, we estimate:

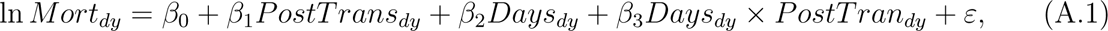

where the outcome variable is the logged fatal AMI counts on day *d* in year *y*, after being demeaned by day-of-week and year to eliminate persistent day-of-week effects and year-to-year variation. The other variables are the same as in Equation (1) in the main text. To be consistent with the main analysis, we use 4 weeks as the bandwidth. The standard errors are corrected for heteroskedasticity.

Table D9 reports the estimated effects at the spring and autumn transition dates. The estimated effects are virtually zero, suggesting that the shifts to and from DST do not cause AMI fatal cases.

**Table D9:** The Effect of DST on AMI Mortality

|  | (1) | (2) |
| --- | --- | --- |
|  | Spring | Autumn |
| PostTrans | -0.00334<br>(0.0199) | 0.0179<br>(0.0248) |

*Notes:* This table presents the estimated effects of DST on AMI mortality based on Equation A.1. The level of observations is at the daily level. The sample size is 400. The bandwidth is 4 weeks before and after the DST transition. The mean number of daily AMI deaths is 80.84 with the standard deviation of 11.69. Robust standard errors, corrected for heteroskedasticity, are reported in the parentheses.
Data source: NVSS MCODE

## E. Medium-term Effects of the Time Adjustments

The event-study model of Equation (7) in the manuscript is:

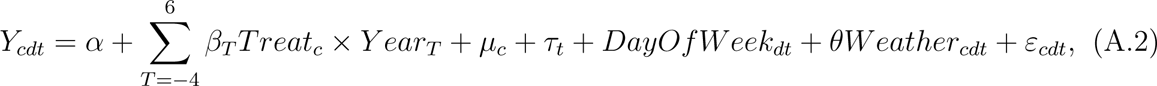

where the additional variable of *Y eart_T_* is an indicator variable for each number of years from the first year of practicing time adjustment, e.g., *T* = 0 in 2006 for most treatment counties. The omitted group is the year before the treatment starts, e.g., *T* = *−*1. The coefficients of interests are *β_T_* ’s, which indicate the changes in the outcome in each year *T* from the reference year of *T* = *−*1 in the treatment counties relative to those changes in the control counties. Thus, we expect the coefficients to be zero for *T <* 0 if we satisfy the parallel trend assumption in the pre-reform period. The standard errors are clustered at the county level.

The results from Equation (A.2) are presented in Figure 3 and discussed in the main text. Importantly, we observe that AMI admission rates are virtually zero up to the year of treatment, ensuring that there is no pre-existing differential trend between the treatment and control counties. The admission rates increase in the first year subject to the spring transition and continues to be positive during the post-reform period.

The other two variables regarding the disease severity do not have statistically significant effects (Online Appendix Table E2).

**Table E1:** The DD Estimates of the Effects of Time Adjustments on AMI

| Variable | (1) | (2) | (3) |
| --- | --- | --- | --- |
| Wk(-3) × PostReform | -0.617<br>(0.654) | -0.620<br>(0.653) | -0.598<br>(0.659) |
| Wk(-2) × PostReform | 0.257<br>(0.587) | 0.270<br>(0.588) | 0.274<br>(0.588) |
| Wk(0) × PostReform | 0.967<br>(0.591) | 0.951<br>(0.588) | 0.960<br>(0.588) |
| Wk(1) × PostReform | 0.888<br>(0.561) | 0.872<br>(0.554) | 0.881<br>(0.554) |
| Wk(2) × PostReform | 0.511<br>(0.524) | 0.500<br>(0.524) | 0.506<br>(0.527) |
| Wk(3) × PostReform | | -0.115<br>(0.600) | -0.261<br>(0.575) |
| Wk(4) × PostReform | | | 0.274<br>(0.755) |
| Mean | 5.084 | 5.084 | 5.084 |
| N | 30,360 | 35,420 | 40,480 |

*Notes:* This table presents the estimated coefficients in Figure 2 in the main text based on Equation (6). All regressions control for the day-of-week effects, daily mean temperature with bins of every 3-degree Celsius, total amount of precipitation, the county fixed effects, the year fixed effects, and the month fixed effects. The level of observations is at the county-daily level. The regressions are weighted by the number of working-age population. The standard errors clustered at the county level are reported in the parenthesis. Column (1) considers the sample of 3 weeks before and after the time transition, and Columns (2) and (3) adds another post-transitional week. The mean values are calculated in the week before the spring transition in the post-reform period.

**Table E2:** The DD Estimates of the Effects of Time Adjustments on AMI

|  | Admission rates | Total charges | Length of stay |
| --- | --- | --- | --- |
|  | (1) | (2) | (3) |
| Treat × PostReform | 0.479*<br>(0.278) | -0.112<br>(0.076) | -0.033<br>(0.024) |
| Mean | 6.223 | 27,978.3 | 4.319 |
| N | 159,160 | 37,026 | 29,929 |
| Fixed effects | Y | Y | Y |
| Controls | Y | Y | Y |

*Notes:* This table presents the effects of the time adjustments on AMI based on the DD model specified by Equation (7) during the DST period. The outcomes are the daily number of hospital admissions per one million population in Column (1), the (log) of total medical charges in Column (2), and the (log) of length of hospital stay in Column (3). To be consistent throughout the paper, the outcomes are first demeaned by daily average temperature with bins of every 3-degree Celsius, total daily precipitation, and day-of-week effects to account for persistent and long-term effects by using all observations throughout the year, whereas including these variables directly into the regressions produces the same results. Treat is a dummy variable for counties that did not practice DST until 2006. The regressions are weighted by the number of working population in Column (1). All regressions include the county fixed effects, the year fixed effects, the month fixed effects. The level of observations are at the daily-county level in Column (1) and at the patient level in Columns (2) and (3). The standard errors are clustered at the county level.
\* $p < 0.10$

## F. Long-term Effects of the Time Adjustments

**Figure F1:**
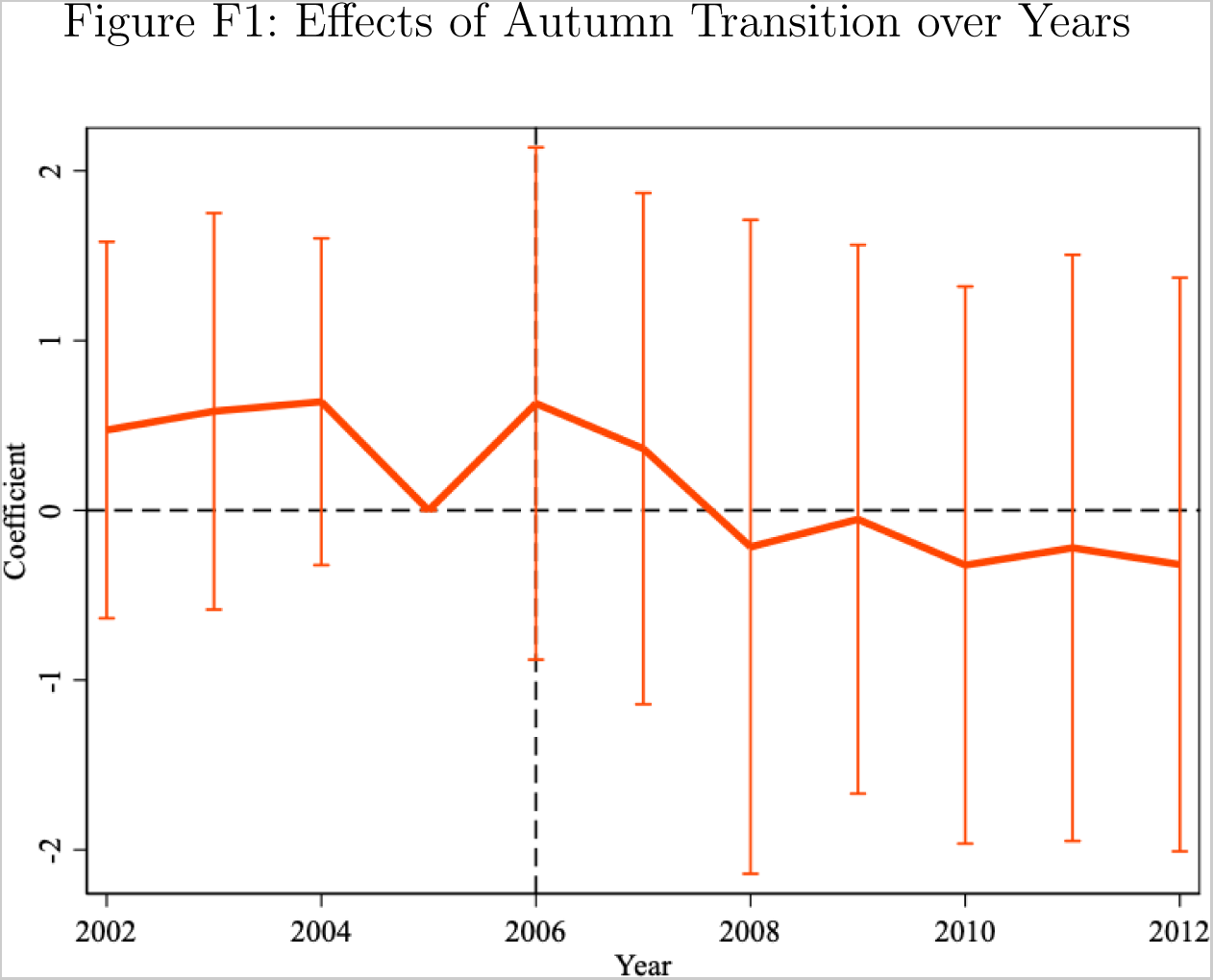
Effects of Autumn Transition over Years *Notes*: This figure presents the effects of the autumn transition on the AMI admission rates per one million working-age population in each year based on the event-study RDDD model specified by Equation (5).

## Footnotes

1 Many countries, such as Russia, Egypt, and Ukraine, have recently abolished DST, and many others, including the EU and U.S., are currently considering terminating its practice. In contrast, Japan is probably the only major economy moving in the other direction, as the government recently announced it was considering reinstating DST.

2 For example, Kotchen and Grant (2011), Kellogg and Wolff (2008), and Sexton and Beatty (2014) find increased electricity usage, whereas others find otherwise (Aries and Newsham 2008; Hill et al. 2010).

3 For instance, studies find that DST affects traffic accidents (Coren 1996; Hicks et al. 1998; Vincent 1998; Coate and Markowitz 2004; Barger et al. 2005; Huang and Levinson 2010; Smith 2016, Bünnings and Schiele 2021), returns in the financial markets (Kamstra et al. 2000; Pinegar 2002; Müller et al. 2009), suicide rates (Berk et al. 2008), criminal activity (Doleac and Sanders 2015), SAT scores (Gaski and Sagarin 2011), and individual well-being (Kountouris and Remoundou 2014).

4 Cardiovascular disease is the leading cause of morbidity and mortality in the U.S. and around the world; in the US., the estimated annual incidence of heart attack was 605,000 new cases and 200,000 recurrent cases between 2005 and 2014, whereas treating heart attacks is one of the most expensive treatments in U.S. (AHA, 2021).

5 Kotchen and Grant (2011) exploit the same setting to examine the relationship between DST and energy consumption in Indiana.

6 Indeed, seasonal variation in AMI occurrence with preponderance in winter is well known in the medical literature (Marchant et al. 1993; Spencer et al. 1998).

7 More detailed and extensive literature is described in Online Appendix A.

8 In contrast, Jin and Ziebarth (2020) find increased sleep time at the autumn transition using the Behavioral Risk Factor Surveillance System in 2013–2016.

9 Online Appendix Table A1 summarizes the existing studies.

10 More details are presented in Online Appendix B.

11 In conjunction with DST shifts, several counties were approved to change the time zones, which often negated the time change.

12 Our sample includes hospital admissions with the diagnosis code of 410 based on ICD-9. The data also report the subcategories at the 5-digit level, indicating that 94.1% of the cases in our data constitute initial episodes. Since our data are discharge records, the diagnosis codes reflect the actual discharge diagnoses rather than an, often tentative, admission diagnosis. Also, since our data are discharge records, our sample does not include fatal cases. We discuss that including fatal cases does not alter our conclusions in Online Appendix D.6.

13 The credibility and completeness of the data are ensured by the Indiana State Department of Health: “IHA [Indiana Hospital Association] processes the records for accuracy, consistency and completeness, and requests resubmissions as necessary” (Indiana Department of Health, 2012).

14 We present analogous evidence using medical charges and length of hospital stay in Online Appendix D.1, for whom DST appears to have little effect, suggesting that the DST-induced AMI are as severe as others.

15 We obtained daily precipitation and temperature information, important factors that change on a daily basis and are likely to be associated with AMI, from NOAA Global Historical Climatology Network Daily (GHCN-D) (NOAA, 2022). In practice, we follow Smith (2016) by first demeaning the outcome by these controls to account for persistent and long-term effects by using all observations throughout the year. Yet, running Equation (3) with controls to account for the effects of these controls specifically around the DST transitions produces the same results.

16 The population data come from the Surveillance, Epidemiology, and End Results Program at National Institute of Health, Single Year of Age County Population Estimates (SEER, 2019).

17 The initial selection of bandwidth at four weeks is set to the one that allows a reasonable comparison with the weekly aggregated analysis (e.g., a multiple of seven days) and is deliberately close to those provided by optimal bandwidth selectors proposed by Ludwig and Miller (2007) (29 days), Imbens and Kalyanaraman (2012) (23 days), and Calonico et al. (2014) (18 days). See Online Appendix D.3 for the estimates based on optimal bandwidths selected by these methods as well as additional bandwidths, which give consistent estimates.

18 Note that while the day-of-week effects are absorbed in the RDDD analysis, to be consistent throughout the paper, we first demean the outcome by the county effects, year effects, day-of-week effects, and weather effects.

19 For ease of interpretation, we remove counties that the time adjustment was offset in 2006 by switching their time zones, yet including these counties has virtually no impact on the results due to their small sample size.

20 Since the number of counties that started practicing DST in 2007 is small, our results are not affected even when we consider a staggered adoption design (de Chaisemartin and D’Haultfœuille 2020), since negative weight is not detected, or when we drop those counties and keep the same year of the policy reform.

21 Online Appendix D.1 provides additional figures for other dependent variables (e.g., total charges and length of hospital stay).

22 Online Appendix D.1 presents analogous results using the RD model. Essentially, the estimated effects based on the RDDD analysis closely mimic the differences in the point estimates between Panel A and B in the RD results for each transitional period.

23 More results by the weekly-level analysis are presented in Online Appendix 2.

24 We present the estimates based on the DD analyses in Online Appendix E.

25 While the precise mechanism to explain the results is unclear, both the American Academy of Sleep Medicine and the Society for Research on Biological Rhythms made recommendations in favor of a permanent shift to Standard Time due primarily to less morning light exposure during DST (Rishi et al. 2020; Roenneberg et al. 2019).

26 Online Appendix F shows that the autumn transition has no impact on the AMI incidence in any particular year.

1 In 2017, the Nobel Prize for medicine was awarded to Jeffrey Hall, Michael Rosbash, and Michael W. Young for their work on the internal clock. Their work revealed that our biological clock helps regulate sleeping, feeding, and hormone production, and any misalignment when we experience “jet lag” can have negative consequences in our health.

2 The peripheral clocks are considered to be similarly affected by external stimuli such as daylight, feeding-fasting cycle, or physical activity and/or to be regulated by the central clock, although the exact mechanisms remain unclear.

3 It is found that 7–9 hours of sleep is ideal for adults, although people in the United States sleep on average about 6.8 hours a day, and people across the contemporary world in general sleep shorter than before. Any shorter and longer duration of sleep contributes to the progression of cardiovascular diseases, including hypertension, coronary artery disease, risk of arrhythmia, stroke, and diabetes mellitus. See the literature summarized in Cundrle et al. (2014).

4 Jin and Ziebarth (2020) find a contradictory result of increased sleep time at the autumn transition using the Behavioral Risk Factor Surveillance System in 2013–2016.

5 The RD normally compares the mean difference between days before and after the DST transitions. In measuring the IR in this study and others in the medical literature, the comparison group is the mean observations of several weeks before and after the DST transitions. Although it can account at least partially for time trends over the transitions, its validity depends crucially on the implicit assumption that AMI that occur in these control weeks after the DST transitions are free from DST effect.

6 The initial idea of DST dates back to Benjamin Franklin in 1789, who proposed that forwarding time by an hour in summer would more effectively utilize the sunshine and save candles.

7 The initial dates of “spring forward” and “fall back” were established on the last Sunday in April and the last Sunday in October, respectively. The 1986 amendment of the Uniform Time Act moved forward the beginning of DST by three weeks. Later, the Energy Policy Act of 2005 further pushed forward the start date by three weeks to the second Sunday in March and pushed backward the end date by one week to the first Sunday in November, effective of 2007.

8 Prior to 1966, Indiana already had internal disputes over the practice of DST between, on the one hand, urban legislators, who were in favor of Eastern Standard Time (EST), and, on the other, their rural counterparts, who were in favor of Central Time Zone (CST). This conflict led Indiana to request the Secretary of Transportation under the Department of Transportation (DOT) to split the state between time zones. The Uniform Time Act of 1966 exempted states with multiple time zones from the practice of DST, and Indiana was one of them. The legislation also allowed individual states to opt out as long as they did so uniformly. For example, Arizona, Hawaii, and Michigan were exempted from observing DST (although Michigan started observing DST later).

9 The northwestern counties include Jasper, Lake, LaPorte, Newton, Porter, and Starke, and the south-western counties include Gibson, Pike, Posey, Spencer, Vanderburgh and Warrick.

10 These include Clark, Floyd, Harrison, Dearborn and Ohio counties.

11 The counties that observed DST up to 2006 include: Clark, Dearborn, Floyd, Gibson, Harrison, Jasper, Lake, La Porte, Newton, Ohio, Porter, Posey, Spencer, Vanderburgh, Warrick.

12 These counties include Daviess, Dubois, Knox, Martin, Perry, Pike, Pulaski, and Starke.

13 The disparity in the results between total charges and length of stay is not clearly explicable. Although in general there is a strong correlation between the two, a large body of evidence has shown that the severity of illness does not explain much variation in the length of hospital stay (Hicks and Kammerling 1993).

14 The percentage change in the expected number of AMI admissions is given by *exp*(*β*) *−* 1.

15 As one can see from Table C1, the average age is 53.1 years old with a relatively low standard deviation of 7.9 years, indicating that there is little dispersion in age. This does not allow us to even look at each 10-year age bin. Thus, we decided to cluster all ages above and below 50, a round number around the mean.

16 The employment rate data come from the American Community Survey (United States Census Bureau, 2012). We use the data in 2012.

17 Analysis of restricted data through the NCHS Research Data Center is approved by the NCHS ERB. The findings and conclusions in this paper are those of the author(s) and do not necessarily represent the views of the Research Data Center, the National Center for Health Statistics, or the Centers for Disease Control and Prevention.

## References

American Heart Association, “2021 Heart Disease and Stroke Statistics Update Fact Sheet: At-a-Glance,” 2021. https://www.heart.org/-/media/phd-files-2/science-news/2/2021-heart-and-stroke-stat-update/2021_heart_disease_and_stroke_statistics_update_fact_sheet_at_a_glance.pdf (accessed June 30, 2022).

Aries, Myriam BC and Guy R Newsham, “Effect of daylight saving time on lighting energy use: A literature review,” Energy policy, 2008, 36 (6), 1858–1866.

Barger, Laura K, Brian E Cade, Najib T Ayas, John W Cronin, Bernard Rosner, Frank E Speizer, and Charles A Czeisler, “Extended work shifts and the risk of motor vehicle crashes among interns,” New England Journal of Medicine, 2005, 352 (2), 125–134.

Barnes, Christopher M. and David T. Wagner, “Changing to Daylight Saving Time Cuts Into Sleep and Increases Workplace Injuries,” Journal of Applied Psychology, 2009, 94 (5), 1305–1317.

Berk, Michael, Seetal Dodd, Karen Hallam, Lesley Berk, John Gleeson, and Margaret Henry, “Small shifts in diurnal rhythms are associated with an increase in suicide: The effect of daylight saving,” Sleep and Biological Rhythms, 2008, 6 (1), 22–25.

Bünnings, Christian and Valentin Schiele, “Spring Forward, Don’t Fall Back: The Effect of Daylight Saving Time on Road Safety,” The Review of Economics and Statistics, 2021, 103 (1).

Burgess, Helen J, “Evening ambient light exposure can reduce circadian phase advances to morning light independent of sleep deprivation,” Journal of sleep research, 2013, 22 (1), 83–88.

Calonico, Sebastian, Matias D Cattaneo, and Rocio Titiunik, “Robust nonparametric confidence intervals for regression-discontinuity designs,” Econometrica, 2014, 82 (6), 2295–2326.

Coate, Douglas and Sara Markowitz, “The effects of daylight and daylight saving time on US pedestrian fatalities and motor vehicle occupant fatalities,” Accident Analysis & Prevention, 2004, 36 (3), 351–357.

Coren, Stanley, “Daylight savings time and traffic accidents,” New England Journal of Medicine, 1996, 334 (14), 924–925.

de Chaisemartin, Clément and Xavier D’Haultfœuille, “Two-Way Fixed Effects Estimators with Heterogeneous Treatment Effects,” American Economic Review, 2020, 110 (9), 2964–2996.

Doleac, Jennifer L and Nicholas J Sanders, “Under the cover of darkness: How ambient light influences criminal activity,” Review of Economics and Statistics, 2015, 97 (5), 1093–1103.

Durgan, David J and Martin E Young, “Relationship Between Myocardial Ischemia/Reperfusion and Time of Day,” in “Translational Cardiology,” Springer, 2012, pp. 1–38.

Gaski, John F and Jeff Sagarin, “Detrimental effects of daylight-saving time on SAT scores.,” Journal of Neuroscience, Psychology, and Economics, 2011, 4 (1), 44.

Hicks, Gregory J, James W Davis, and Robert A Hicks, “Fatal alcohol-related traffic crashes increase subsequent to changes to and from daylight savings time,” Perceptual and motor skills, 1998, 86 (3), 879–882.

Hill, Simon I., Frédéric Desobry, Elizabeth W. Garnsey, and Yu-Foong Chong, “The impact on energy consumption of daylight saving clock changes,” Energy Policy, 2010, 38, 4955–4965.

Huang, Arthur and David Levinson, “The effects of daylight saving time on vehicle crashes in Minnesota,” Journal of safety research, 2010, 41 (6), 513–520.

Imbens, Guido and Karthik Kalyanaraman, “Optimal bandwidth choice for the regression discontinuity estimator,” The Review of Economic Studies, 2012, 79 (3), 933–959.

Imbens, Guido and Thomas Lemieux, “The Regression Discontinuity Design: Theory and Applications,” Journal of Econometrics, 2008, 142 (2), 611–850.

Indiana Department of Health, “Hopsital Discharge Data,” 2012. https://www.in.gov/health/oda/data-analysis-and-risk-factors/hospital-discharge-data/indiana-hospital-discharge-data-documentation/.

Jin, Lawrence and Nicolas R. Ziebarth, “Sleep, Health, and Human Capital: Evidence from Daylight Saving Time,” Journal of Economic Behavior and Organization, 2020.

Kamstra, Mark J, Lisa A Kramer, and Maurice D Levi, “Losing sleep at the market: The daylight saving anomaly,” American Economic Review, 2000, 90 (4), 1005–1011.

Kellogg, Ryan and Hendrik Wolff, “Daylight time and energy: Evidence from an Australian experiment,” Journal of Environmental Economics and Management, 2008, 56 (3), 207–220.

Kotchen, Matthew J and Laura E Grant, “Does daylight saving time save energy? Evidence from a natural experiment in Indiana,” Review of Economics and Statistics, 2011, 93 (4), 1172–1185.

Kountouris, Yiannis and Kyriaki Remoundou, “About time: Daylight saving time transition and individual well-being,” Economics Letters, 2014, 122 (1), 100–103.

Ludwig, Jens and Douglas L Miller, “Does Head Start improve children’s life chances? Evidence from a regression discontinuity design,” The Quarterly journal of economics, 2007, 122 (1), 159–208.

Manfredini, Roberto, Fabio Fabbian, Rosaria Cappadona, Alfredo De Giorgi, Francesca Bravi, Tiziano Carradori, Maria Elena Flacco, and Lamberto Manzoli, “Daylight Saving Time and Acute Myocardial Infarction: A Meta-Analysis,” Journal of Climinical Medicine, 2019, 8 (404).

Marchant, Bradley, Kulasegaram Ranjadayalan, Robert Stevenson, Paul Wilkinson, and Adam D Timmis, “Circadian and seasonal factors in the pathogenesis of acute myocardial infarction: the influence of environmental temperature,” British Heart Journal, 1993, 69 (5), 385–387.

Müller, Luisa, Dirk Schiereck, Marc W Simpson, and Christian Voigt, “Daylight saving effect,” Journal of Multinational Financial Management, 2009, 19 (2), 127–138.

NEWSRT, “Time stops: Russia abolishes daylight saving time practice,” 2011.

NOAA, “Global Historical Climatology Network Daily,” 2022. https://docs.opendata.aws/noaa-ghcn-pds/readme.html (accessed on February 6, 2022).

Pinegar, J Michael, “Losing sleep at the market: Comment,” American Economic Review, 2002, 92 (4), 1251–1256.

Rishi, Muhammad Adeel, Omer Ahmed, Jairo H. Barrantes Perez, Michael Berneking, Joseph Dombrowsky, Erin E. Flynn-Evans, Vicente Santiago, Shannon S. Sullivan, Raghu Upender, Kin Yuen, Fariha Abbasi-Feinberg, R. Nisha Aurora, Kelly A. Carden, Douglas B. Kirsch, David A. Kristo, Raman K. Malhotra, Jennifer L. Martin, Eric J. Olson, Kannan Ramar, Carol L. Rosen, James A. Rowley, Anita V. Shelgikar, and Indira Gurubhagavatula, “Daylight saving time: an American Academy of Sleep Medicine position statement.,” Journal of Clinical Sleep Medicine, 2020, 16, 1781–1784.

Roenneberg, Till, Anna Wirz-Justice, Debra J. Skene, Sonia Ancoli-Israel, Kenneth P. Wright, Derk-Jan Dijk, Phyllis Zee, Michael R. Gorman, Eva C. Winnebeck, and Elizabeth B. Klerman, “Why should we abolish daylight saving time?,” Journal of Biological Rhythms, 2019, 34, 227–230.

Sexton, Alison L. and Timothy K. M. Beatty, “Behavioral responses to Daylight Savings Time,” Journal of Economic Behavior and Organization, 2014, 107 (A), 290–307.

Smith, Austin C., “Spring Forward at Your Own Risk: Daylight Saving Time and Fatal Vehicle Crashes,” American Economic Journal: Applied Economics, 2016, 8 (2), 65–91.

Spencer, Frederick A., Robert J. Goldberg, Richard C. Becker, and Joel M. Gore, “Seasonal Distribution of Acute Myocardial Infarction in the Second National Registry of Myocardial Infarction,” Journal of the American College of Cardiology, 1998, 31 (6), 1226– 1233.

Surveillance, Epidemiology, and End Results Program, “U.S. County Population Data - 1969-2019,” 2019. https://seer.cancer.gov/popdata.thru.2019/download.html#single (accessed on February 5, 2022).

Vincent, Alex, “Effects of daylight savings time on collision rates.,” The New England Journal of Medicine, 1998, 339 (16), 1167.

Witte, D.R., D.E. Grobbee, M.L. Bots, and A.W. Hoes, “A meta-analysis of excess cardiac mortality on monday,” European Journal of Epidemiology, 2005, 20, 401–406.

## References

Berson, David M, Felice A Dunn, and Motoharu Takao, “Phototransduction by retinal ganglion cells that set the circadian clock,” Science, 2002, 295 (5557), 1070–1073.

Centers for Disease Control and Prevention, “National Vital Statistics System Multiple Cause of Death,” 2011. https://www.cdc.gov/nchs/nvss/index.htm.

Čulić, Viktor, “Daylight saving time transitions and acute myocardial infarction,” Chronobiology International, 2013, 30 (5), 662–668.

Cundrle, Ivan, Andrew D Calvin, and Virend K Somers, “Sleep deprivation and the cardiovascular system,” in “Sleep deprivation and disease,” Springer, 2014, pp. 131–147.

Cunningham, Timothy J., Anne G. Wheaton, ES Ford, and Janet B. Croft, “Racial ethnic disparities in self-reported short sleep duration among US-born and foreign-born adults,” Ethnicity & Health, 2016, 21 (6), 628–638.

Department of Transportation, “Standard Time Zone Boundary in the State of Indiana,” 2006.

Dranove, David, Daniel Kessler, Mark McClellan, and Mark Satterthwaite, “In More Information Better? The Effects of ‘Report Cards’ on Health Care Providers,” Journal of Political Economy, 2003, 111 (3).

Durgan, David J and Martin E Young, “Relationship Between Myocardial Ischemia/Reperfusion and Time of Day,” in “Translational Cardiology,” Springer, 2012, pp. 1–38.

Gelman, Andrew and Guido Imbens, “Why High-Order Polynomials Should Not Be Used in Regression Discontinuity Designs,” Journal of Business and Economic Statistics, 2019, 37 (3), 447–456.

Giuntella, Osea and Fabrizio Mazzonna, “Sunset time and the economic effects of social jetlag: evidence from US time zone borders,” Journal of Health Economics, 2019, 65, 210–226.

Harrison, Yvonne, “The impact of daylight saving time on sleep and related behaviours,” Sleep medicine reviews, 2013, 17 (4), 285–292.

Hicks, N. and R. Kammerling, “The relationship between a severity of illness indicator and mortality and length-of-stay,” Health Trends, 1993, 25 (2), 65–68.

Janszky, Imre and Rickard Ljung, “Shifts to and from daylight saving time and incidence of myocardial infarction,” New England Journal of Medicine, 2008, 359 (18), 1966–1968.

Janszky, Imre, Staffan Ahnve, Rickard Ljung, Kenneth J Mukamal, Shiva Gautam, Lars Wallentin, and Ulf Stenestrand, “Daylight saving time shifts and incidence of acute myocardial infarction–Swedish Register of Information and Knowledge About Swedish Heart Intensive Care Admissions (RIKS-HIA),” Sleep medicine, 2012, 13 (3), 237–242.

Jiddou, Monica R, Mark Pica, Judy Boura, Lihua Qu, and Barry A Franklin, “Incidence of myocardial infarction with shifts to and from daylight savings time,” The American journal of cardiology, 2013, 111 (5), 631–635.

Kantermann, Thomas, Myriam Juda, Martha Merrow, and Till Roenneberg, “The human circadian clock’s seasonal adjustment is disrupted by daylight saving time,” Current Biology, 2007, 17 (22), 1996–2000.

Kirchberger, I., K. Wolf, M. Heier, B. Kuch, W. von Scheidt, A. Peters, and C. Meisinger, “Are daylight saving time transitions associated with changes in myocardial infarction incidence? Results from the German MONICA/KORA Myocardial Infarction Registry,” BMC Public Health, 2015, 15 (778).

Lahti, Tuuli A, Sami Leppäamäki, Jouko Lönnqvist, and Timo Partonen, “Transition to daylight saving time reduces sleep duration plus sleep efficiency of the deprived sleep,” Neuroscience letters, 2006, 406 (3), 174–177.

Monk, Timothy H and Simon Folkard, “Adjusting to the changes to and from Daylight Saving Time,” Nature, 1976, 261 (5562), 688.

Portaluppi, Francesco, Ruana Tiseo, Michael H Smolensky, Ramón C Hermida, Diana E Ayala, and Fabio Fabbian, “Circadian rhythms and cardiovascular health,” Sleep medicine reviews, 2012, 16 (2), 151–166.

Sipilä, Jussi O.T., Päivi Rautava, and Ville Kytö, “Association of daylight saving time transitions with incidence and in-hospital mortality of myocardial infarction in Finland,” Annals of Medicine, 2016, 48 (1-2), 10–16.

Smolensky, Michael H, Ramon C Hermida, Richard J Castriotta, and Francesco Portaluppi, “Role of sleep-wake cycle on blood pressure circadian rhythms and hypertension,” Sleep medicine, 2007, 8 (6), 668–680.

Takeda, Norihiko and Koji Maemura, “Circadian clock and cardiovascular disease,” Journal of cardiology, 2011, 57 (3), 249–256.

Tonetti, Lorenzo, Alex Erbacci, Marco Fabbri, Monica Martoni, and Vincenzo Natale, “Effects of transitions into and out of daylight saving time on the quality of the sleep/wake cycle: an actigraphic study in healthy university students,” Chronobiology international, 2013, 30 (10), 1218–1222.

Toro, Weily, Robson Tigre, and Breno Sampaio, “Daylight Saving Time and incidence of myocardial infarction: Evidence from a regression discontinuity design,” Economics Letters, 2015, 136, 1–4.

United States Census Bureau, “American Community Survey,” 2012. https://www.census.gov/programs-surveys/acs (accessed on July 25, 2019.

Watson, Nathaniel F., Safwan Badr, Gregory Belenky, Donald L. Bliwise, Orfeu M. Buxton, Daniel Buysse, David F. Dinges, James Gangwisch, Michael A. Grandner, Clete Kushida, and, et al, “Recommended Amount of Sleep for a Healthy Adult: A Joint Consensus Statement of the American Academy of Sleep Medicine and Sleep Research Society,” Journal of Clinical Sleep Medicine, 2015, 11 (6), 591–592.

